# Exome sequencing of 628,388 individuals identifies common and rare variant associations with clonal hematopoiesis phenotypes

**DOI:** 10.1101/2021.12.29.21268342

**Authors:** Michael D. Kessler, Amy Damask, Sean O’Keeffe, Michael Van Meter, Nilanjana Banerjee, Stefan Semrau, Dadong Li, Kyoko Watanabe, Julie Horowitz, Yariv Houvras, Christopher Gillies, Joelle Mbatchou, Ryan R. White, Jack A. Kosmicki, Michelle G. LeBlanc, Marcus Jones, Regeneron Genetics Center, GHS-RGC DiscovEHR Collaboration, David J. Glass, Luca A. Lotta, Michael N. Cantor, Gurinder S. Atwal, Adam E. Locke, Manuel A. R. Ferreira, Raquel Deering, Charles Paulding, Alan R. Shuldiner, Gavin Thurston, Will Salerno, Jeffrey G. Reid, John D. Overton, Jonathan Marchini, Hyun M. Kang, Aris Baras, Gonçalo R. Abecasis, Eric Jorgenson

## Abstract

Clonal hematopoiesis (CH) refers to the expansion of certain blood cell lineages and has been associated with aging and adverse health outcomes. Here, we use exome sequence data on 628,388 individuals to identify 40,208 carriers of clonal hematopoiesis of indeterminate potential (CHIP). Using genome-wide and exome-wide association analyses, we identify 27 loci (24 novel) where germline genetic variation influences CH/CHIP predisposition, including missense variants in the DNA-repair gene *PARP1* and the lymphocytic antigen coding gene *LY75* that are associated with reduced incidence of CH/CHIP. Analysis of 5,194 health traits from the UK Biobank (UKB) found relationships between CHIP and severe COVID outcomes, cardiovascular disease, hematologic traits, malignancy, smoking, obesity, infection, and all-cause mortality. Longitudinal analyses revealed that one of the CHIP subtypes, *DNMT3A*-CHIP, is associated with the subsequent development of myeloid but not lymphoid leukemias, and with solid cancers including prostate and lung. Additionally, contrary to previous findings from the initial 50,000 UKB exomes, our results in the full sample do not support a role for IL-6 inhibition in reducing the risk of cardiovascular disease among CHIP carriers. Our findings demonstrate that CHIP represents a complex set of heterogenous phenotypes with shared and unique germline genetic causes and varied clinical implications.

## Introduction

As humans age, somatic alterations accrue in the DNA of hematopoietic stem cells (HSCs) due to mitotic errors and DNA damage. Alterations that convey a selective growth advantage can lead to expansion of particular cell lineages, a phenomenon known as clonal hematopoiesis (CH). The presence of CH has been associated with an increased risk of hematological neoplasms, cytopenias, cardiovascular disease, infection, and all-cause mortality^1–5^. For this reason, identifying germline causes of CH has the potential to inform our understanding of initiating events in the development of these common diseases.

Large-scale studies of the germline causes of CH have utilized samples from the UK Biobank (UKB) and other large cohorts, but those studies have been mostly limited to CH phenotypes that can be assessed using SNP array genotype data, such as mosaic chromosomal alternations (mCA) and mosaic loss of sex chromosomes (mLOX and mLOY)^4,6,7^. Identifying subjects with clonal hematopoiesis of indeterminate potential (CHIP), which is defined by somatic protein-altering mutations within genes that are recurrently mutated in CH, requires sequencing of blood^1,2^. Once a clone has expanded sufficiently, the somatic variants from this clone can be captured along with germline variation through exome sequencing. Exome sequencing also enables the detection of rare variation that has a large impact on phenotypes, which can further elucidate critical genes and pathways and potential therapeutic targeting^8,9^. To date, the largest genetic association study of CHIP included 3,381 CHIP mutation carriers in a sample of 65,405 individuals and identified four common variant loci^10^.

Here, we use exome sequencing data to characterize CHIP status across 454,803 UK Biobank^9^ and 173,585 Geisinger MyCode Community Health Initiative (GHS) participants. We then conduct a common variant genome-wide association study (GWAS) and rare variant and gene burden exome-wide association study (ExWAS) of CHIP across 27,331 CHIP mutation carriers from UKB. We perform replication in 12,877 CHIP mutation carriers from the GHS cohort. To identify germline predictors of specific CH driver mutations, we conduct genetic association analyses of carriers of CHIP mutations from individual CHIP genes. We then compare genetic association findings for CHIP to those from analyses of other CH phenotypes determined from somatic alterations in the blood, including mCA, mLOX, mLOY, and telomere length. While prior GWAS studies of these non-CHIP CH phenotypes have been conducted^4,6,11^, none have evaluated the impact of very rare variants. Therefore, the ExWAS we perform here represents the first systematic large-scale exploration of the impact of rare variants on the genetic susceptibility of these phenotypes. Finally, we examine the clinical consequences of somatic CHIP mutations and germline predictors of CHIP in several ways. We first conduct a PheWAS^12^ of germline predictors of CHIP to understand their biological functions, and then test cross-sectional phenotype associations of CHIP carrier status across 5,194 traits in the UKB. We then test the risk of incident cancer, cardiovascular disease, and all-cause mortality among specific CHIP-gene mutation carriers.

## Results

### Calling CHIP

We used exome sequencing data from 454,803 and 173,585 individuals from the UK Biobank (UKB) and Geisinger Health System MyCode Community Health Initiative cohort (GHS) cohorts, respectively, to generate the largest callsets of CHIP carrier status to date (see Methods). Briefly, we called somatic mutations using Mutect2 in a pipeline that included custom QC filtering (Figure 1A), and ultimately restricted our analysis to 23 well defined and recurrent CHIP-associated genes. This focused analysis identified 29,669 variants across 27,331 individuals in UKB (6%), and 14,766 variants across 12,877 individuals in GHS (7.4%). *DNMT3A*, *TET2*, *ASXL1*, *PPM1D*, and *TP53* were the most commonly mutated genes in both cohorts (Figure S1A). While the GHS cohort has a wider age range, and therefore a larger number of older individuals, the prevalence by age was similar across cohorts, and reached ~15% by age 75 (Figure 1B, C). Prevalence of CHIP-gene-specific mutations was consistent with recurrence patterns, with mutations in the most commonly mutated CHIP genes increasing in prevalence at younger ages (Figure 1D, E). In older GHS individuals, we were able to see some an exception to this shared pattern, as the prevalence of *ASXL1* mutations appears to taper off at older ages (Figure 1E).

**Figure 1.**
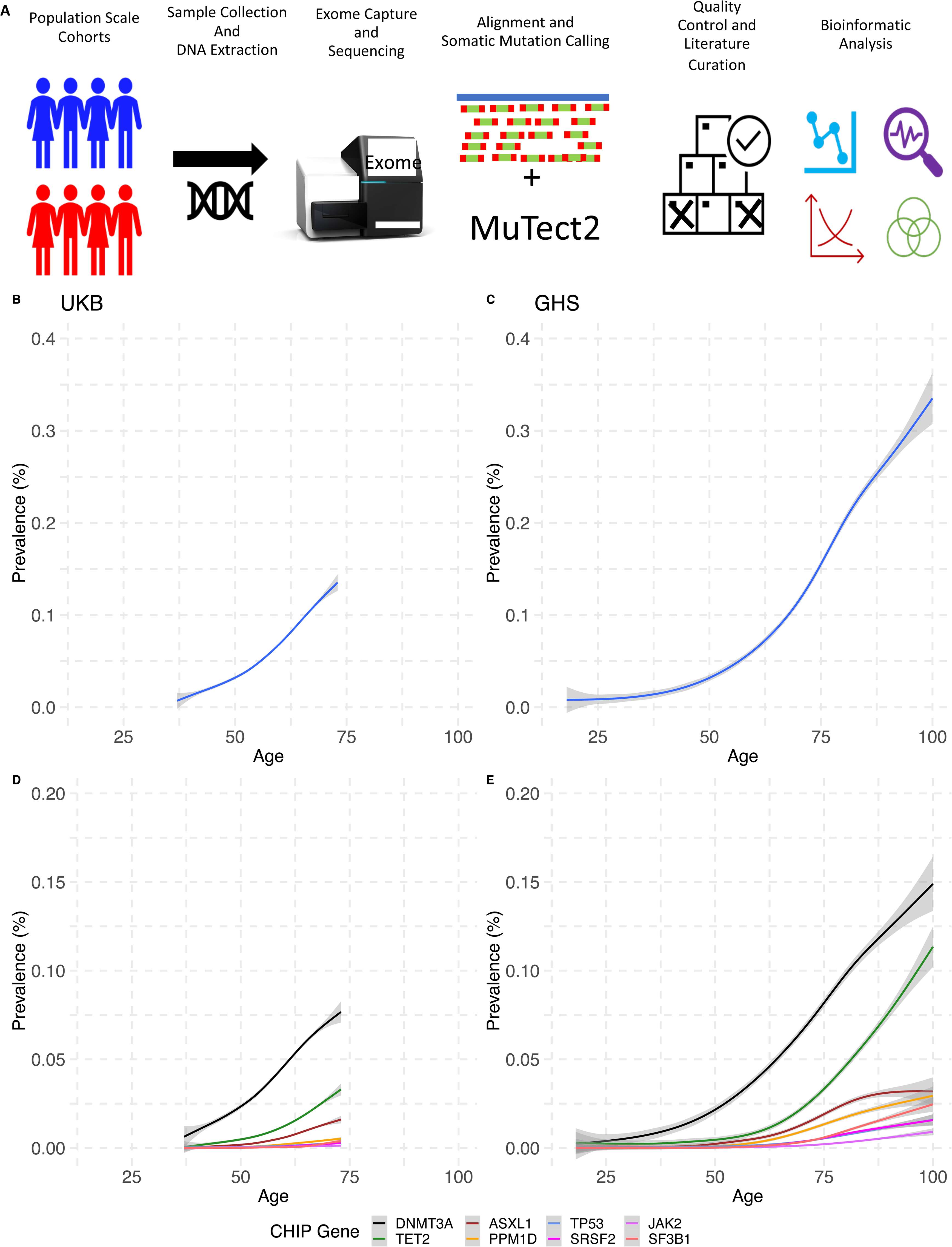
Prevalence and Count Distribution of Clonal Hematopoiesis of Indeterminate Potential (CHIP) Gene Mutations and Common Variant Genome-Wide Association Results. A. Total number of individuals with mutations (y axis, log_10_ scale) in each of the 23 genes that were used to determine CHIP status across the UKB (blue) and GHS (red) CHIP callsets. B. Splines showing the prevalence of CHIP mutations by age in the UKB and GHS cohorts. C. Similar to B, splines showing the prevalence of CHIP mutations by age in the UKB and GHS cohorts, but broken down by CHIP gene for each of the top-8 most common CHIP genes. D. Pairwise mutation counts across the UKB (left) and DiscoverEHR (right) callsets across individuals with at least two identified CHIP mutations. The color scale reflects the significance of the P-value for enrichment of the presence of pairs of gene mutations as determined by Fisher’s exact test. P-values are –log_10_ transformed, and values > 25 were set to the color corresponding to 25 for visualization (see Table S3 for complete enrichment results).

Somatic mutations in certain CHIP genes co-occurred in the same individual more frequently than expected, with particularly strong enrichments found for individuals having either *SRSF2*, *JAK2*, or *SF3B1* mutations along with *TET2* (Figure S1B,C, Table S1). Other enriched CHIP pairs found across both the UKB and GHS callsets included *DNMT3A* and *TET2, TET2* and *ASXL1, DNMT3A* and *PPM1D, IDH2* and *SRSF2*, *IDH2* and *ASXL1*, *TET2* and *CALR*, and *TET2* and *ETNK1*. Among CHIP mutation carriers, 11,125 (40%) had at least one CHIP somatic mutation at a high variant allele fraction (VAF > 10%) in the UKB (5,981 [47%] had high VAF CHIP in GHS). Except for *JAK2*, which had a relatively high average VAF, the more common CHIP mutations generally had lower VAFs than the rarer and more canonically leukemogenic mutations within individuals with a single CHIP mutation (Figure S2A). Amongst the eight most recurrently mutated CHIP genes, average age among carriers with mutations in a single CHIP gene was lowest in *DNMT3A* carriers (59.95) and highest in *SRSF2* (62.72) and *SF3B1* carriers (63.51).

Among individuals with multiple CHIP mutations, *JAK2* mutations consistently had the highest VAF (Figure S2B). Maximum VAF among all CHIP carriers increased linearly with the number of CHIP mutations identified in an individual (beta = 0.101, P < 1 × 10^−16^, Figure S2C), consistent with the concept of accumulation of multiple mutations during progressive clonal expansion. Telomere length was not substantially different across CHIP mutation carriers (Figure S2D). Although restricting the analysis to individuals with high VAF reduced available sample size and analytic power (since high VAF represents further progressed and expanded CHIP), clinical associations with CHIP have typically been reported for high VAF CHIP carriers^2,131^, and so we generally assessed all identified CHIP carriers first and then also evaluated high VAF CHIP carriers on their own.

### CHIP demographics

Compared with controls, CHIP carriers in both the UKB and GHS cohorts were older and more likely to be heavy smokers, consistent with previous studies^10^ (Tables S2, S3). While our cohorts are predominantly comprised of European ancestry individuals, prevalence of CHIP was similar across ancestries (Figure S3). In multivariate logistic regression models, each additional year of age was strongly associated with an increased risk of CHIP in UKB (OR = 1.08 [1.077-1.082], P < 10^−300^) and GHS (OR = 1.06 [1.061-1.065], P < 10^−300^), and heavy smoking was strongly associated with CHIP carrier status in both UKB (OR = 1.17 [1.135-1.209], P = 3.00 × 10^−23^) and GHS (OR = 1.22 [1.12-1.34], P = 9.6 × 10^−6^). Overall, our results suggest that the prevalence of CHIP will double every 9-12 years of life. These associations with age and smoking were stronger when restricting to high VAF CHIP carriers. In our multivariate modeling, women were significantly more likely to be CHIP mutation carriers than men in UKB (OR = 1.08 [1.05-1.11], P = 6.4 × 10^−7^), but not in GHS (OR = 1.02 [0.95-1.08, P = 0.62]). Overall, these associations were consistent when restricting to high VAF CHIP carriers, although the risk of high VAF CHIP was not significantly greater in women in UKB (OR = 1.035 [0.99-1.08], P = 0.126).

### Genetic association analyses of CHIP mutation carrier status

We first conducted genetic association analyses in the UKB cohort to identify germline loci associated with the risk of developing CHIP. In the common variant (MAF > 0.5%) GWAS, which included 25,657 cases and 342,866 controls of European ancestry, we identified 27 loci (24 novel) harboring 57 independently associated variants (Figure 2, Table S4). To confirm these signals, we conducted a replication analysis in 9,523 CHIP cases and 105,502 controls of European ancestry from the GHS cohort. Of the 57 independent signals, 53 had directionally consistent effects, 14 of the 27 sentinel variants at the associated loci were nominally significant (at P < 0.05, only 1.35 variants were expected to show consistent association), and 5 were significant at a Bonferroni level of significance (at P < 0.0019, only 0.05 variants were expected to show consistent association, Table S5).

**Figure 2.**
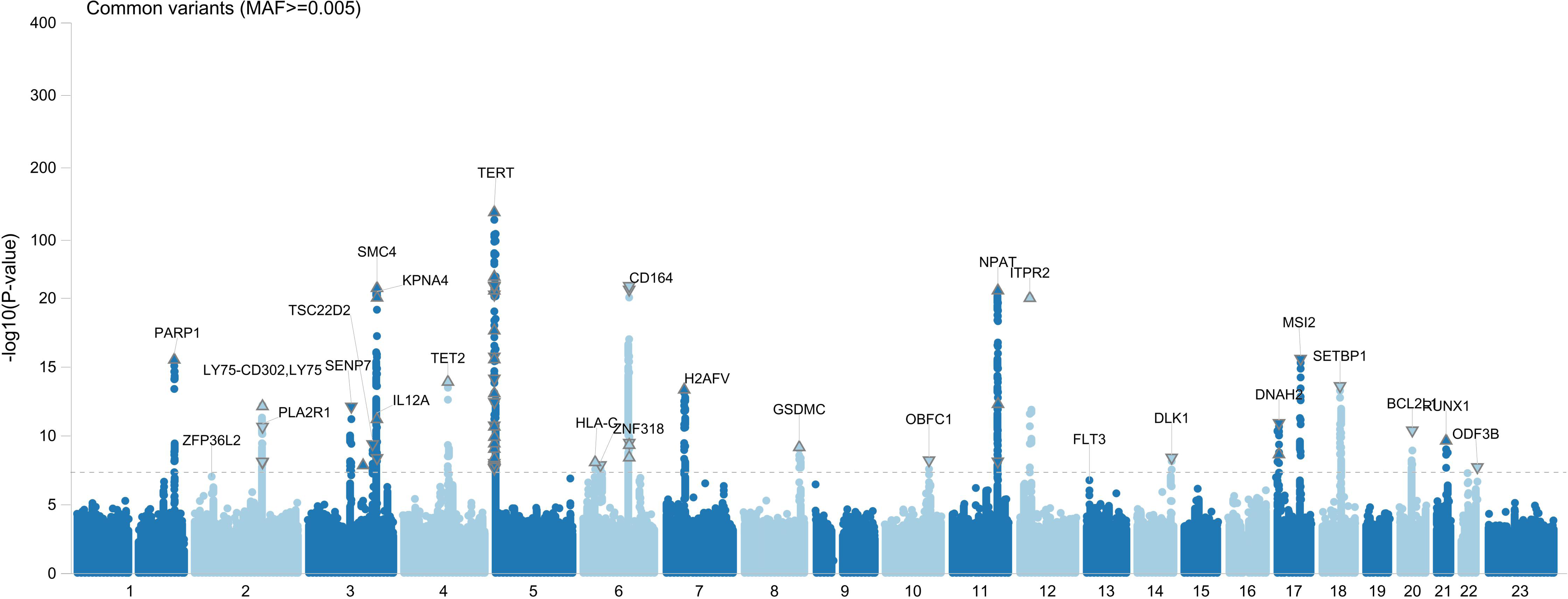
GWAS of Clonal Hematopoiesis of Indeterminate Potential (CHIP) Manhattan plot showing results from a genome-wide association analysis of CHIP. 27 loci reach genome-wide significance (P <= 5e-8, dashed line), and top-associated variants per locus are labeled with their nearest gene.

Since the CHIP phenotype we constructed is based on the presence of rare variants in recurrently mutated genes and since distinguishing somatic and germline variants is challenging, even after exome sequencing, rare variant and gene burden associations from genome-wide analysis will sometimes feature the same variants and genes through which the phenotype is defined. Furthermore, other associated rare variants (i.e., those which were not used to condition the CHIP phenotypes) may themselves be somatic variants, which achieve higher VAFs during hematopoietic clonal expansion and become indistinguishable from germline variants by best practice variant calling. To avoid circularity, we exclude all such variants from our reported results. Further, we assessed whether significantly associated rare variants had low variant allele fractions (VAFs) across carriers, as well as whether an individual’s age-at-sample-collection was associated with variant carrier status (as both these tests suggest a somatic origin for a variant). For genome-wide significant rare variant and gene burden associations for which we had exome data, we reported these VAF and age-association results along with genetic association results in order to provide resolution as to whether such associations are likely driven by germline or somatic variation.

We identified 20 rare variants across 3 loci with significant CHIP associations (P <= 7.14 × 10^−10^, Table S6) and five genes with significant burden associations (P <= 3.6 × 10^−7^, Table S7). All 3 rare variant loci overlapped with common variant loci and therefore likely represent germline associations. These loci were found at the end of chromosome 2, at the *SMC4* locus on chromosome 3, and at a locus on chromosome 22 that overlaps *CHEK2*. In GHS, 18 of the 20 rare variants had directionally consistent effects, 16 of those were significant at P < 0.05 (1 expected), and 13 were significant at P < 0.0025 (0.05/20; 0.05 expected). The genes associated via burden testing include two cancer-associated genes *ATM* and *CHEK2* involved in DNA repair, the telomere maintenance and DNA replication associated gene *CTC1*, the *CLPTM1L* gene that has been associated with resistance to apoptosis in tumors, and the Notch pathway and cancer associated gene *KDELC2*. All 5 genes had directionally consistent effects in GHS, and 2 (*ATM* and *CHEK2*) were significant at P < 0.05, and VAF and age-association calculations suggest that all are driven by germline variation (Table S7).

Even though our power to detect associations in non-EUR populations was limited, we ran genome-wide association analyses for African ancestry individuals, South Asian ancestry individuals, and East Asian ancestry individuals. Although no associations reached genome-wide significance, 15/57 showed directionally consistent effects across all populations (Table S8; 7/57 expected). We also attempted to replicate the recently described CHIP-risk-increasing association reported for the African ancestry-specific *TET2* enhancer variant (rs144418061-A)^10^, but did not find any significant association in African ancestry individuals (OR = 1.38 [0.87-2.19], P = 0.174, AAF = 0.036), nor in European ancestry individuals, among whom we identified 90 heterozygous carriers (OR = 1.02, [0.45-2.31], P = 0.97, AAF = 0.000014).

For each index variant associated with CHIP, we queried its associations across 1,557 binary and quantitative health traits from UKB for which we had previously performed genetic association analysis^9^ (Table S9). Overall, traits with significant associations predominantly consisted of blood measures (i.e. cells counts and biomarker levels), size and impedance measures, and auto-immune phenotypes. Four SNPs (rs9264393-A, rs80093687-T, rs78378222-G, rs2275652-C) featured numerous significant phenotypic associations across these traits (Figure S3).

### Individual CHIP gene mutation carrier association analyses

To identify CHIP-subtype specific risk variants, we defined gene-specific CHIP phenotypes for each of the eight most commonly mutated CHIP genes. For each subtype, we selected individuals with mutations in one gene of the eight genes and no mutations in any of the other genes used to define CHIP. We then conducted association analyses comparing these single CHIP gene carriers to CHIP-free controls, and observed shared, unique, and opposing effects of associated loci on CHIP subtypes (Figure 3, Tables S10-S26). We also replicated these CHIP-subtype-specific associations in the GHS cohort. Despite many of the rarest variants from UKB not being present in GHS, and having limited power overall, we replicated the majority of sentinel signals at the 0.05 level, and directions of effect were consistent across > 90% of signals. When variants were associated with multiple CHIP subtypes, there effect sizes were generally only modestly correlated (r2 < 0.2; Figure S5).

**Figure 3.**
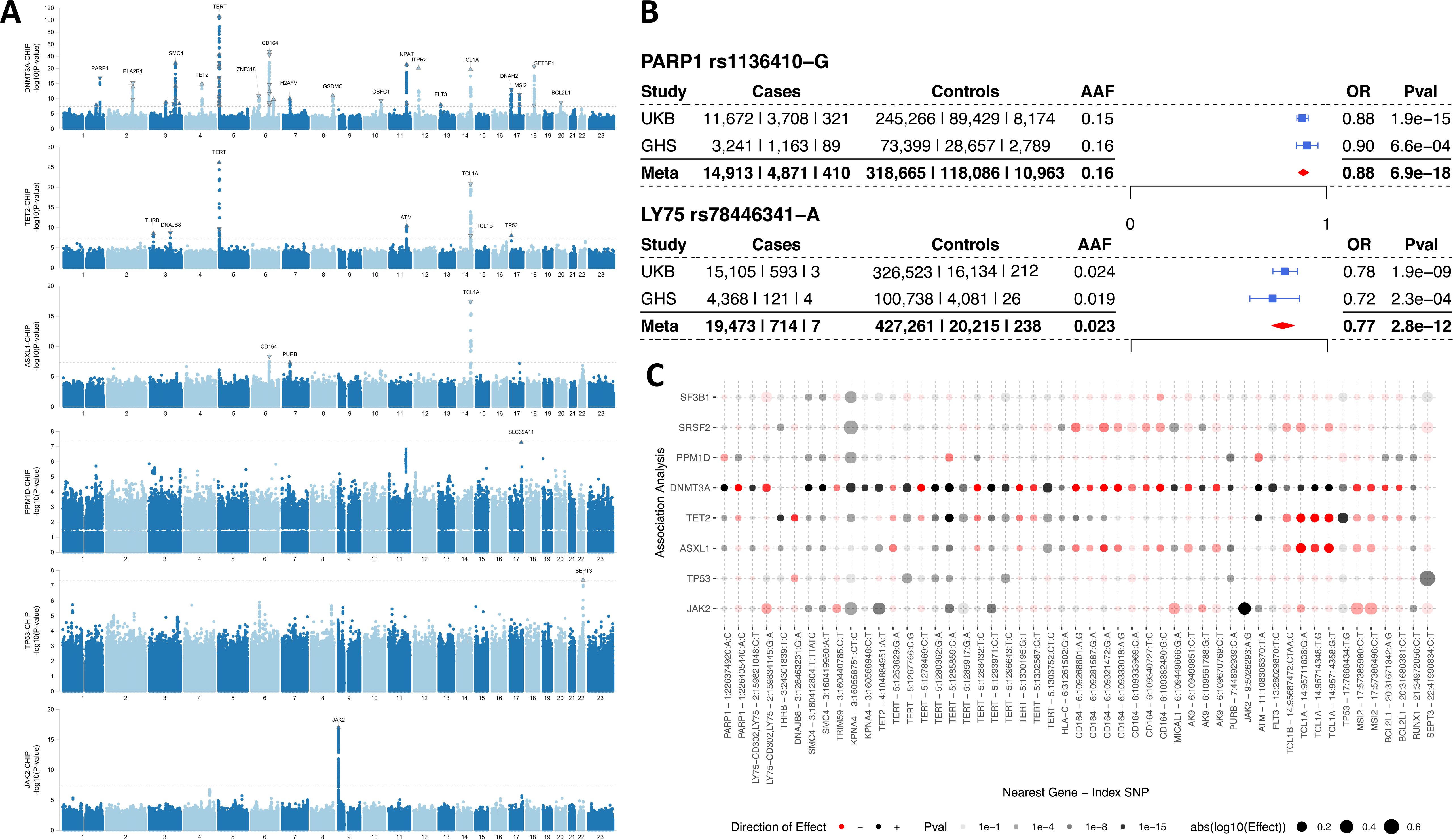
Genome-wide Association Results for Carriers of Single CHIP Gene Mutations and Age Distributions by Gene. A. Manhattan plots from analyses of single CHIP gene mutation carrier status, compared to clonal-hematopoiesis-free controls, are shown for the six genes that had at least one genome wide significant common variant association. B. Table and forest plots showing associations with *PARP1* and *LY75* protective missense variants in UKB, GHS, and a meta-analysis of the two. C. Pleiotropy plot comparing effect size estimates across CHIP gene specific association analyses for all loci showing genome wide significant association with CHIP overall or any CHIP gene specific phenotype.

*DNMT3A*, which was the most commonly mutated gene in the CHIP, had the largest number of significantly associated loci (n = 23), most of which overlapped with the overall CHIP association signals – suggesting that germline variants in these CHIP genes can substitute for somatic mutations in predisposing to clonal expansions. Although most loci harbored variants that increased CHIP risk, two exceptions are noteworthy. At a locus in chromosome 1, rs1136410 (Val762Ala in *PARP1*) was associated with reduced risk of *DNMT3A*-CHIP (OR = 0.88 [0.85-0.91], P = 1.88 × 10^−15^). At a locus on chromosome 2, rs78446341 (Pro1247Leu in *LY75*) was also associated with reduced risk of *DNMT3A*-CHIP (OR = 0.78 [0.72-0.84], P = 3.70 × 10^−10^). The former is an eQTL that is estimated to reduce the risk of *PARP1* expression (P = 7.20 × 10^−17^)^14^. *PARP1* plays a role in DNA damage repair, whereas *LY75* is thought to be involved in regulating the proliferation of B lymphocytes. Both signals replicated in GHS (Figure 3B).

Among loci associated with multiple CHIP subtypes, we observed genome-wide significant association signals at the *TCL1A* locus that were not present in the overall CHIP analysis. This locus is notable because it exhibited opposing effects across CHIP subtypes (Figure 3D, Table S27), with lead SNPs (e.g. rs2887399-T, rs11846938-G, rs2296311-A) at the locus associated with an increased risk of *DNMT3A*-CHIP (OR = 1.14[1.11-1.17], P = 1.8 × 10^−20^) but a reduced risk of *TET2*-CHIP (OR = 0.76[0.72-0.80], P = 3.8 × 10^−21^) and *ASXL1*-CHIP (OR = 0.70[0.65-0.76], P = 6.9 × 10^−18^). Effect estimates from the other five CHIP-gene specific association analyses were also consistent with protective effects (Figure 3D, Table S27). This is consistent with findings from a recent genetic association study of CHIP in the TOPMed cohort, which identified a genome-wide significant positive association of the *TCL1A* locus and *DNMT3A*-CHIP as well as a nominally significant opposing signal for *TET2*-CHIP^10^. Additionally, the *DNMT3A*-CHIP increasing allele has been found to reduce the risk of mLOY in a recent GWAS^6^. This observation suggests that *DNMT3A*-CHIP is distinct among clonal hematopoietic subtypes with regard to the genetic influence of the *TCL1A* locus, which may relate to the fact that TCL1A and DNMT3A both play key roles in DNA methylation^16^.

Beyond the *TCL1A* locus, the gene-specific CHIP analyses also identified patterns of effects that distinguish CHIP subtypes (Figure 3C, Table S27). For example, in comparison to other CHIP subtypes, associated loci often showed opposing effects on *PPM1D*-CHIP, notably at the *PARP1* locus on chromosome 1, the *LY75* locus on chromosome 2, the *TERT* locus on chromosome 5, the *ATM* locus on chromosome 11, and the *BCL2L1* locus on chromosome 20. In contrast to the discordant effects at these loci, signals at the *SMC4* locus on chromosome 3 (which includes the *TRIM59* and *KPNA4* genes) were strongly consistent across all CHIP subtypes. As reported by others^17^, the *JAK2* 46/1 locus was strongly associated with an increased risk of *JAK2*-CHIP (OR = 2.24 [1.86-2.69], P = 9.22 × 10^−18^).

Rare variant results for *DNMT3A*-CHIP were similar to those from CHIP overall, with significant associations observed at the *SMC4* and *CHEK2* loci (Table S11), and significant gene burden associations observed at *ATM, CHEK2, CTC1, KDELC2* and *CLPTM1L* (Table S12). We found two significant rare variant associations with *TP53*-CHIP (P <= 7.14 × 10^−10^, Table S21), including a risk increasing missense variant (rs754853158-T) in the *NEURL4* gene (OR = 1284 [242-6830], P = 4.63 × 10^−17^). Other rare variant associations were only nominally significant (Tables S11-S21). One of these consisted of a rare loss-of-function stop gain mutation in the *PML* gene that associated with *TP53*-CHIP (OR = 276 [37-2062], P = 4.26 × 10^−8^), and was noteworthy given that *PML* has a well described association with promyelocytic leukemia, is known to be a tumor suppressing transcription factor, and has been reported to interact directly with TP53^18,19^. Another noteworthy association was between rare loss of function (and missense) variants in the *NFE2* gene (gene burden framework) and *JAK2*-CHIP in UKB (OR = 141 [23-865], P = 8.81 × 10^−8^, Table S25) and *DNMT3A*-CHIP in GHS (OR = 11.09 [4.44-27.71], P = 2.58 * 10^−7^), which provides support for *NFE2* gene function loss as a driver of clonal hematopoiesis.

### Genetic comparisons between CHIP, mosaic chromosomal alterations, and telomere length

To evaluate the relationship between CHIP and other forms of somatic alterations of the blood, we utilized phenotype information on other types of clonal hematopoiesis and on telomere length that are available on UKB participants^4,6,7,11^. We first evaluated the phenotypic overlap between CHIP and mLOY, mLOX, and autosomal mosaic chromosomal alterations (mCAaut). CHIP is distinct from mCA phenotypes (mCAaut, mLOX, and mLOY), with >80% of CHIP carriers having no identified mCAs (Figure 4A). However, CHIP carriers are enriched for mCAs (OR = 1.43, P = 2.14 × 10^−101^). Carriers of only a single CH phenotype (i.e. CHIP, mLOY, mLOX, or mCAaut) were younger on average than those with multiple CH lesions, and mCAaut and CHIP carriers were youngest among single CH phenotype carriers (Figure 4B). The fact that mLOY occurs in older individuals but is also relatively common suggests that the processes driving and/or following such clonal genetic loss happen more quickly than do other somatic alterations (and/or that loss of Y is often recurrent and polyclonal).

**Figure 4.**
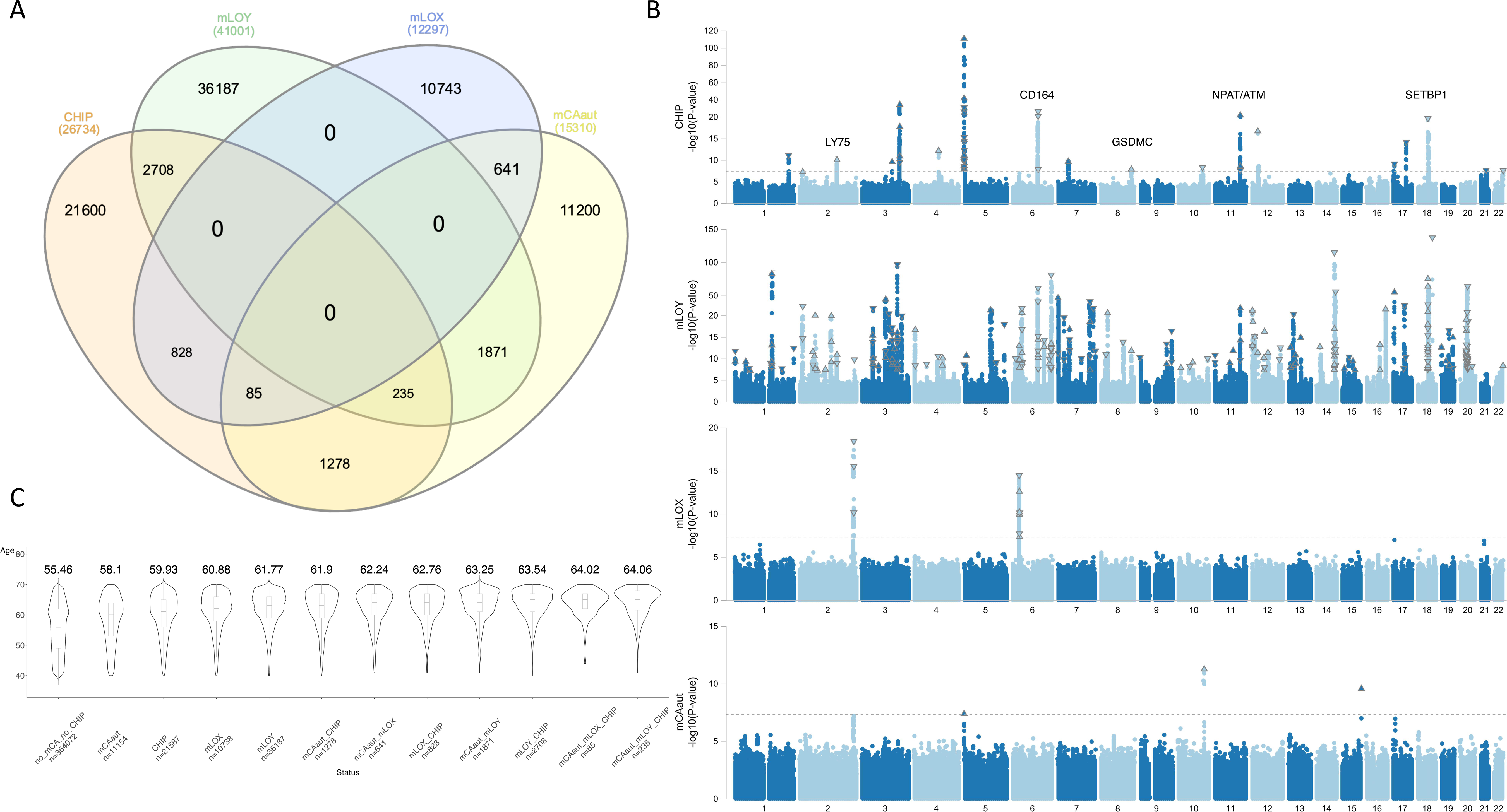
Comparisons Between CHIP and Somatic Clonal Hematopoietic Phenotypes Defined by Chromosomal Alteration. A. Venn diagram showing the overlap between CHIP and other somatic mosaic alteration phenotypes across samples from UKB. B. Manhattan plots of results from association analyses of exclusive CHIP and mCA phenotypes (i.e. carriers have only CHIP or an mCA). The majority of associated loci are found in association with CHIP and/or mLOY (labels reflect the nearest gene(s) at loci associated with both CHIP and mLOY). C. Violin plots of the age distributions of carriers of CHIP and/or mCA phenotypes (label = distributional mean). Individuals without any CHIP or mCA are the youngest, and individuals with an increasing number of somatic lesion are older on average.

We then conducted GWAS and ExWAS analyses of these somatic alteration phenotypes and evaluated the shared germline genetic contributions to CHIP and these traits. Genome-wide genetic correlation^20,21^ was nominally significant between CHIP and mLOY, the two most heritable of these somatic alteration-based traits (r_g_ = 0.27, P = 0.014, uncorrected, Table S28). We then evaluated whether common genetic variants associated with any of the CH phenotypes described above were shared across traits (Figure 4, Tables S29-S36). When comparing effect sizes across index variants from loci found to be genome-wide significant in any of our CH phenotype associations, we observed significant correlations between CHIP and mLOY (Beta = 5.0, R^2^ = 0.263, P < 3 × 10^−24^) and mLOX (Beta = 1.1, R^2^ = 0.242, P =31 × 10^−22^), but not with mCAaut (Beta = 0.061, R^2^ = 0.005, P = 0.2) (Figure S6). Notably, variants at 4 loci (marked by the nearest genes *NPAT/ATM, LY75, CD164,* and *GSDMC*) showed similar associations with both CHIP and mLOY, while variants at the *SETBP1* locus were negatively associated with CHIP and positively associated with mLOY. mLOY and mLOX also show significant positive correlation across these genetic loci (Beta = 1.7, R^2^ = 0.168, P = 3 × 10^−15^), whereas mCAaut is not correlated with either mLOY (Beta = −0.17, R^2^ = 0.001, P = 0.5) or mLOX (Beta = 0.05, R^2^ = 0.001, P = 0.5). On the whole, these analyses suggest that despite being distinct clonal hematopoietic phenotypes, CHIP and mLOY and mLOX share multiple germline genetic risk factors.

### Exome analyses identified novel genes involved in mCAaut, mLOX, mLOY, and telomere length

While the common variant association analyses of other somatic alteration phenotypes were undertaken for the purpose of comparing to CHIP, and our results are consistent with recent published associations for these non-CHIP UKB somatic alteration phenotypes^4,6,7^, we also identified novel rare variant and gene burden associations via ExWAS analyses (Tables S29-S39). Notably, we found a novel risk reducing association between a rare missense variant in the *KNTC1* gene (rs61751321-T, AAF = 0.003, L317F) and the mLOY phenotype (OR = 0.58[0.48-0.70], P = 7.7 × 10^−9^). We also found a rare missense variant in the *TIE1* gene (rs140190628-T, AAF = 3.9 × 10^−4^, S470L) that increases the risk of acquiring the mCAaut phenotype (OR = 8.5[5.6-13.0], P = 4.5 × 10^−23^), as well as a rare missense variant in the *PTPRF* gene (rs142117073-C, AAF = 8.56 × 10^−5^, Y1621H) that also increases mCAaut risk (16.47 [6.07-44.74], P = 3.89 × 10^−8^). We also identified a rare frameshift variant in the *TM2D3* gene (rs770752834-GT, AAF = 6.0 × 10^−5^) that is strongly associated with increased risk of mCAaut (34.25 [11.99-97.85], P = 4.16 × 10^−11^), which underscores the previously described associations between the *TM2D3* gene disruption and clonal hematopoiesis^6,7^.

Gene burden testing identified a signal supporting a recently described^22^ risk increasing association between rare loss of function variants in the *GIGYF1* gene and mLOY (6.59 [3.32-13.06], P = 6.71 × 10^−8^), and a risk increasing association between rare loss of function variants in the *RC3H1* gene and mCAaut (5.60 [3.34-9.39], P = 5.96 × 10^−11^). Burden tests also support the *PRR14L* gene as a novel CH gene (13.40 [8.51-21.09, P = 3.42 × 10^−29^), as VAF and age calculations suggest that the association with rare loss of function variants in this gene is highly likely to be driven by somatic variation (Table S34).

Finally, identified rare variant associations with telomere length (LTL) include high effect size missense and/or nonsense variants in genes associated with telomere biology (*TERF1, POT1, NAF1, ACD, SAMHD1, HBB, RTEL1,* and *TINF2*), and gene burden associations suggesting that loss of function of the *DCLRE1B* gene significantly increases telomere length (0.61 [0.47-0.74], P = 2.57 × 10^−19^) and loss of function of the *PARN* gene significantly decreases telomere length (−0.61 [−0.73 − −0.49], P = 8.18 × 10^−23^) (Table S36, S37). A significant positive gene burden association between rare loss of function variants in the *CTC1* gene and telomere length is interesting given that this gene has been reported to be involved in multiple aspects of telomere maintenance^23,24^, and suggests that disruption of *CTC1* has a net elongating effect on telomeres.

### Phenotypic associations with CHIP and CHIP-associated variants

Clonal hematopoiesis has been associated with increased risk of hematologic malignancy and cardiovascular disease, as well as other health outcomes including all-cause mortality, susceptibility to infection^3,4,25,26^. To test for expected as well as potentially novel associations, we performed cross-sectional association analyses across 4,871 traits (2,459 binary traits and 2,421 quantitative traits) available from the UK Biobank and curated as part of our efforts for the UKB Exome Sequencing Consortium. We performed Firth-type penalized logistic regression using CHIP gene mutation carrier status as the binary outcome for each of the 23 CHIP genes in our callset, with age, sex, and ten genetic principal components as covariates. Our results are consistent with previous findings, with the majority of associated phenotypes deriving from cardiovascular, hematologic, neoplastic, infectious, renal, and/or smoking-related causes (Figure 5, S7, Tables S40).

**Figure 5.**
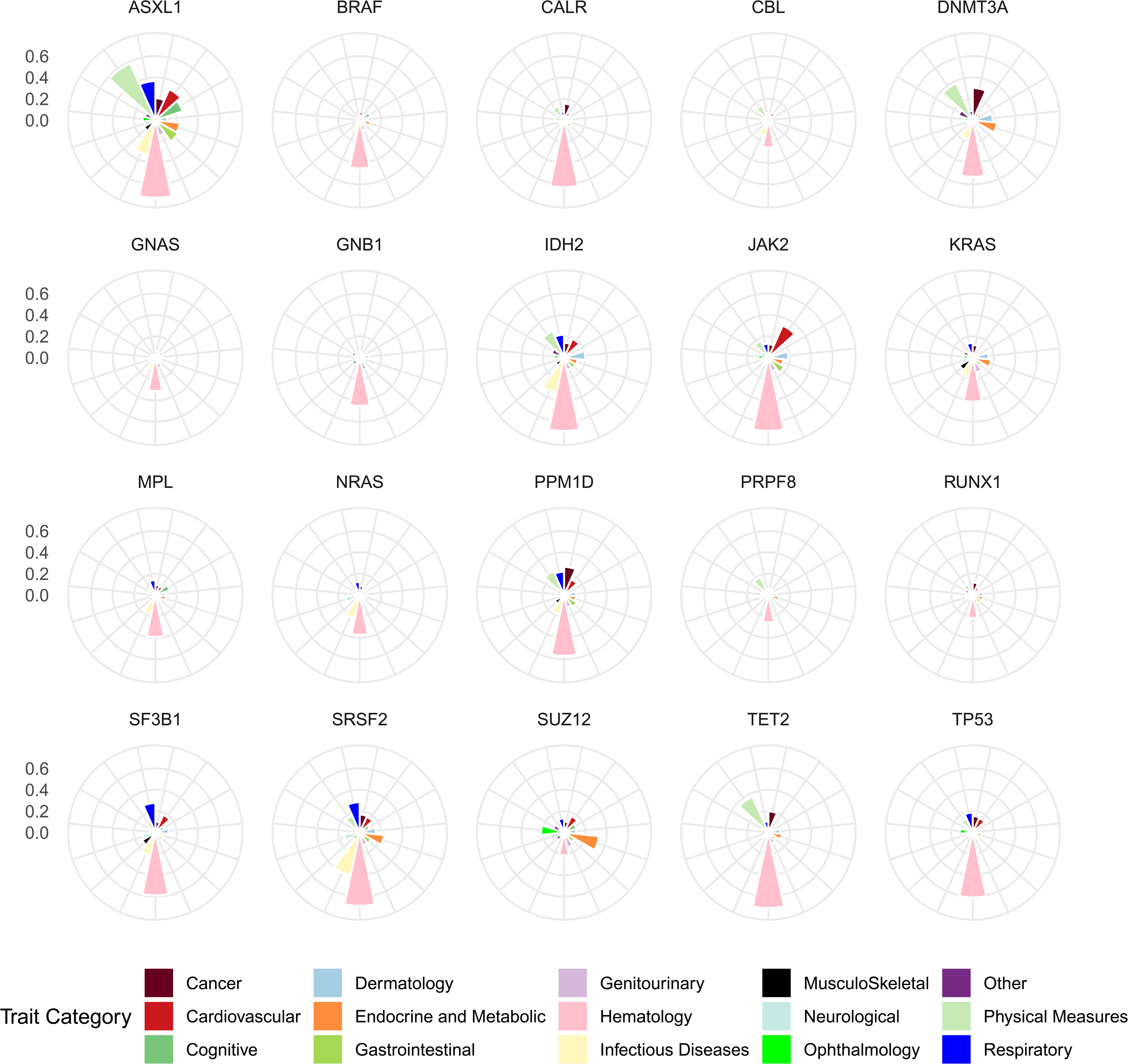
Phenome Association Profiles Per CHIP Subtype. Profiles are shown for each CHIP gene subtype reflecting phenome-wide association results. The y-axis (concentric circles) represents the proportion of phenotypes within a trait category that were nominally associated (P <= 0.05) with carrier status of the CHIP gene. A CHIP gene had to have at least one disease category with the proportion of associated phenotypes >= 0.2 in order to be included in the figure. As expected, hematological traits show the largest proportion of phenotypic trait associations overall. The largest number of cancer associations are seen for DNMT3A-CHIP, whereas JAK2-CHIP shows the highest proportion of cardiovascular associations. Respiratory associations are most pronounced for ASXL1-CHIP. SUZ12-CHIP shows a unique profile across CHIP subtypes, with a higher proportion of ophthalmological and endocrine associations.

*ASXL1*-CHIP was associated with the largest number and widest range of traits, although many of these associations traced to correlates of smoking. *SUZ12*-CHIP showed a distinct association profile amongst CHIP-genes, with a larger proportion of associations in endocrine and ophthalmologic traits than other CHIP-genes. Numerous traits show associations with *DNMT3A*-CHIP and *TET2*-CHIP that were in opposing directions, including white blood cell count, platelet count, and neutrophil count, which were all positively associated with *DNMT3A*-CHIP and negatively associated with *TET2*-CHIP. This is not unexpected if *DNMT3A*-CHIP and *TET2-*CHIP are associated with expansion of different subsets of blood cells. Interestingly, body mass index and fat percentage were negatively associated with *DNMT3A*-CHIP and other leukemogenic CHIP mutations (e.g. *JAK2, CALR, MPL*), but are positively associated with other CHIP subtypes (e.g. *TET2* and *ASXL1*). We also observed significant associations between *JAK2* mutations and gout, which fits with increased red blood cell turnover leading to increased uric acid production.

Given recent reports that CH is associated with an increased risk of COVID-19 and other infections^4,27^, we also tested for association between CHIP and COVID-19 infection using data for individuals from the UKB cohort^28^. When restricting to CHIP carriers with VAF > 10%, we found that CHIP carrier status was significantly associated with COVID-19 hospitalization (OR = 1.37 [1.07-1.72], P = 8.7 × 10^−4^) and severe COVID-19 infection (OR = 1.87 [1.28-2.63], P = 6.3 × 10^−4^) in logistic regression models that excluded individuals with any blood cancers and that adjusted for age, sex, smoking, and 5 genetic principal components. In an analysis among CHIP mutation carriers, CHIP VAF (as a quantitative trait) was also associated with COVID-19 hospitalization (OR = 1.29 [1.07-1.55], P = 6.9 × 10^−3^) and severe COVID-19 infection (OR = 1.56 [1.14-2.14], P = 6.0 × 10^−3^).

### Longitudinal analysis of hematological and oncological risk within individuals with CHIP

Given the confounding that can bias cross-sectional association analyses, we performed survival analyses to evaluate whether individuals with CHIP at the time of enrollment and blood sampling in UKB were at an elevated risk of subsequently developing cardiovascular disease, cancer, and all-cause mortality. To do this, we generated aggregate longitudinal phenotypes of cardiovascular disease (CVD), lymphoid cancer, myeloid cancer, lung cancer, breast cancer, prostate cancer, colon cancer, and overall survival (i.e. Any Death). Because prior longitudinal studies of CHIP and the risk of many of these outcomes have focused on high VAF CHIP, we focused on CHIP carriers with VAF >= 0.10 for these analyses.

We observed a significantly elevated risk of CVD in CHIP carriers (HR = 1.10 [1.03-1.17], P = 6.6 * 10^−3^* 10^−8^, Figure S8A), which was driven by *TET2*-CHIP (HR = 1.20 [1.07-1.34], P = 1.7 * 10^−3^, Figure S8C). However, this risk estimate is lower than the HR of 1.59 recently reported by Bick et al.^13^ in an analysis of CHIP from the first 50k UKB participants with exome sequencing data available. Therefore, we restricted our analysis to the 50k individuals from the previous study and found that the estimated HR is indeed higher in this subset (HR = 1.30 [1.06-1.59], P = 0.013, Figure S8B). Bick et al. also observed a cardio-protective effect of *IL6R* rs2228145-C (a genetic proxy for IL-6 inhibition) among CHIP carriers in the 50k UKB subset, so we repeated that analysis in both the 50k UKB subset and full UKB cohort (n = 430,924 in these analyses). We observed the same CHIP-specific protective *IL6R* effect in the 50k subset as previously reported (HR = 0.60 [0.40-0.89], P = 0.012), however we did not find any *IL6R* effect in the full cohort (HR = 0.97 [0.84-1.11], P = 0.64, n = 430,924, Figure 6). These results were consistent when varying which CHIP mutations in our callset we used to define CHIP, as well as when using different VAF thresholds and a variety of CVD endpoint composites (see Methods).

**Figure 6.**
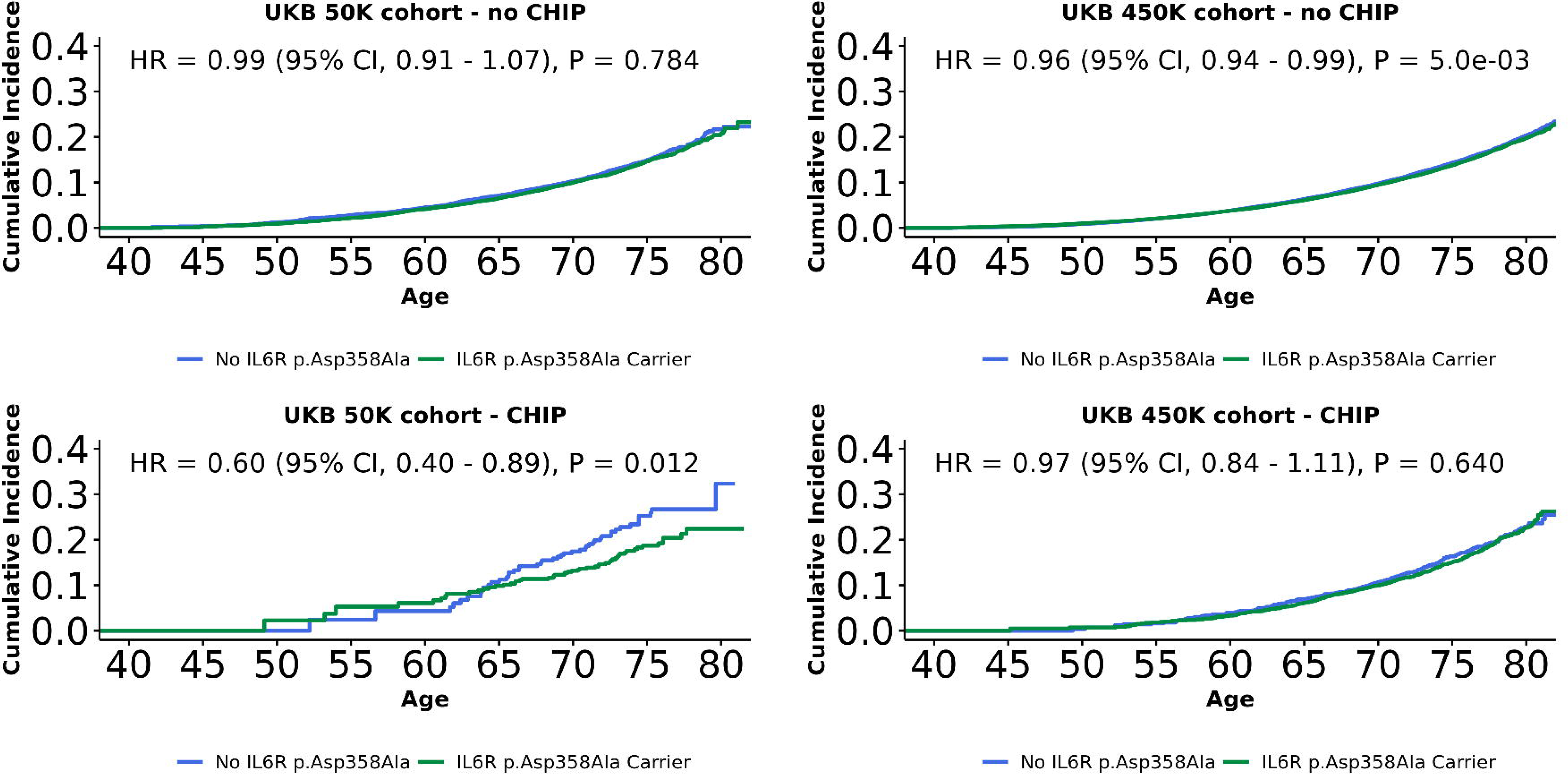
CVD Incidence in IL6R Mutation Carriers with and without CHIP. Survival curves are drawn showing that IL6R p.Asp358Ala mutation carriers are not an elevated risk of CVD incidence (y-axis) in individuals without CHIP in either the first 50K individuals from UKB (A) or the full 450K cohort (B). In contrast, IL6R p.Asp358Ala mutation carriers are estimated to be at a reduced risk of CVD events (CHR = 0.60), but only in the first 50K samples from UKB (C).

We next tested whether CHIP carriers are at an increased risk of hematologic and solid cancers, and whether risk differed by CHIP mutational subtype for the three most common CHIP genes (i.e. *DNMT3A, TET2, ASXL1*, Figure 7). To control for the possibility that toxic chemotherapeutic treatment for previous cancers might drive the development of CHIP mutations^29^, and/or otherwise confound association analyses, we performed all analyses after excluding individuals with any diagnoses of cancer prior to DNA collection. As expected, we found CHIP carriers with VAF >= 0.10 to be at a significantly elevated risk of developing any blood cancer (HR = 3.88 [3.46-4.36], P = 9.10 * 10^−117^). *TET2* mutation carriers were at the greatest risk of developing blood cancers (HR = 4.72 [3.83-5.83], P = 1.70 * 10^−47^), whereas *DNMT3A* mutation carriers had much more modest risk of acquiring blood cancers (HR = 1.70 [1.37-2.11], P = 1.50 * 10^−6^) unless they also had at least one additional CHIP mutation (HR = 3.66 [2.33-5.75], P = 1.70 * 10^−8^, Figure 7A). Subdividing blood cancers into myeloid and lymphoid subtypes, we estimated that high VAF CHIP carriers were at a significantly higher risk of developing myeloid cancers (HR = 7.10 [6.21-8.12], P = 9.30 * 10^−180^, Figure 7B) compared with lymphoid cancers (HR = 1.58 [1.26-1.99], P = 9.30 * 10^−5^, Figure 7C). Furthermore, we estimate that *DNMT3A* mutations did not predispose to lymphoid cancers (HR = 0.93 [0.63-1.37], P = 0.71, Figure 7C).

**Figure 7.**
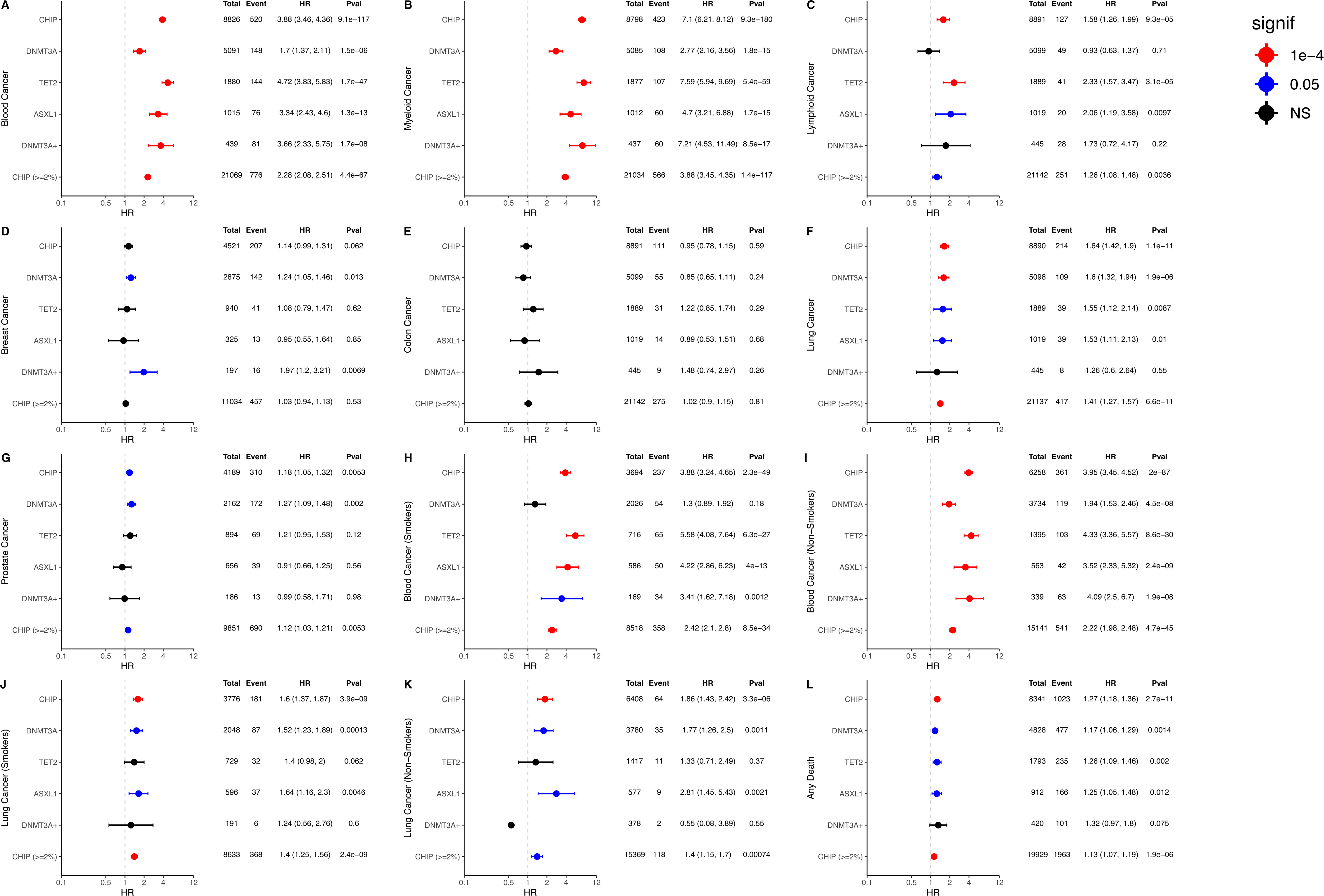
Forest Plots of Hazard Ratios After the Removal of Individuals with Prior Cancers. Forest plots and tables featuring hazard ratio estimates from cox-proportion hazard models are shown. CHIP and its most common subtypes are significantly associated with blood (A) and myeloid (B) cancers, but DNMT3A-CHIP is not associated with lymphoid cancers (C). CHIP is also associated with Breast (D) and Prostate (G) cancers, and with lung cancer (F) in both smokers (K) and non-smokers (L). Here, results from analyses are depicted after the removal of samples that had a diagnosis of malignant cancer prior to sequencing collection.

We then tested whether CHIP carriers were at an elevated risk of developing solid tumors (Figure 7D-G) and found that high VAF carriers are at significantly elevated risk of developing lung cancer (HR = 1.64 [1.42-1.90], P = 1.10 * 10^−11^), modestly increased risk of developing prostate cancer (HR = 1.18 [1.05-1.32], P = 5.30 * 10^−3^), and nominally increased risk of developing breast cancer (HR = 1.14 [0.99-1.31], P = 0.062). We did not find any increased risk for the development of colon cancer (HR = 0.95 [0.78-1.15], P = 0.59). Models estimating event risk on the basis of CHIP mutational subtype (e.g. *DNMT3A*-CHIP) suggest that these associations with prostate and breast cancer are driven primarily by *DNMT3A* mutations (Figure 7).

Given the strong associations between CHIP and both blood and lung cancers, and the associations between smoking and both CHIP and lung cancer, we performed additional analyses stratified by smoking status to test whether these associations were driven by smoking and merely marked by CHIP mutations. High VAF CHIP carriers were at an elevated risk of developing blood cancers in both smokers (HR = 3.88 [3.24-4.65], P = 2.30 * 10^−49^) and non-smokers (HR = 3.95 [3.45-4.52], P = 2.0 * 10^−87^). Notably, lung cancer risk for high VAF CHIP carriers (compared with healthy controls) was significantly elevated among both smokers and non-smokers, and is in fact higher in non-smokers (HR_non-smokers_ = 1.86 [1.43-2.42], P = 3.30 * 10^−6^, HR_smokers_ = 1.60 [1.37-1.87], P = 3.90 * 10^−9^, Figure 7J-K). These associations were driven by *DNMT3A* and *ASXL1* CHIP carriers, with both estimated to have elevated lung cancer risk in both smokers and non-smokers. Overall, these models suggest that CHIP mutation carriers are at an elevated risk of both blood cancer and lung cancer independent of smoking, but that CHIP is likely also marking additional blood cancer risk that results from smoking.

Finally, the risk of death from any cause was significantly elevated among high VAF CHIP carriers (1.27 [1.18-1.36], P = 2.70 × 10^−11^), and was similar across *DNMT3A, TET2*, and *ASXL1* CHIP subtypes (Figure 7L).

### DNMT3A Polygenic Risk Score (PRS) predicts cancers and CHIP subtypes

To further evaluate the relationship between CHIP and these cardiovascular and oncologic phenotypes, we generated a polygenic risk score (PRS) for *DNMT3A*-CHIP, which was largest and best powered CHIP subtype for such an analysis. In multivariate logistic regression models (Table S42), our results were consistent with those from our longitudinal analyses, with *DNMT3A* risk scores significantly positively associated with myeloid cancer (OR = 1.17 [1.13-1.20], P = 4.6 × 10^−22^), lung cancer (OR = 1.09 [1.06-1.12], P = 1.5 × 10^−9^), and prostate cancer (OR = 1.03 [1.01-1.05], P = 6.4 × 10^−4^). Notably, this risk score did not associate with CVD (OR = 0.99 [0.98-1.0], P = 0.11) or with risk of death from any cause (OR = 1.0 [0.99-1.01], P = 0.62). However, in addition to predicting *DNMT3A*-CHIP carrier status (OR = 1.38 [1.36-1.41], P < 10^−300^), this *DNMT3A*-derived risk score was highly associated with being a carrier of *TET2*-CHIP (OR = 1.17 [1.13-1.20], P = 2.4 × 10^−26^), *ASXL1*-CHIP (OR = 1.20 [1.15-1.25], P = 2.8 × 10^−18^), or *JAK2*-CHIP (OR = 1.48 [1.31-1.67], P = 3.5 × 10^−10^).

## Discussion

In this study, we generated the two largest callsets to date of individuals with CHIP mutation carrier information, and we used those calls to identify many novel common and rare variant loci associated with CHIP and CHIP subtypes. These loci, which have shared, unique, and opposing effects on the risk of developing different types of CHIP and other somatic alterations of the blood, highlight the fact that CHIP encapsulates a complex set of heterogeneous phenotypes. We further showed that the genetic etiology of CHIP is reflected in its clinical consequences, as the risk of various clinical conditions was differentially associated across CHIP gene mutations.

The new loci identified in this study provide a foundation on which to investigate the biological mechanisms that lead to specific features of CHIP. For example, among CHIP-associated loci, variants in the *TCL1A* locus that are associated with an increase in the risk of *DNMT3A*-CHIP have the opposite effect on the risk of all other CHIP and CH subtypes. Coupled with recent findings that link *TCL1A*’s role in mLOY to lymphocytes (e.g. B cells)^6^, our results further suggest *TCL1A* as a critical mediator of CH, as well as of CH subtype-specific differences.

Several of the novel loci associated with *DNMT3A*-CHIP harbor genes that are potential targets for the development of new treatments to prevent or slow the expansion of CHIP clones. Both *PARP1* and *LY75* have missense variants that are associated with a reduced risk of *DNMT3A*-CHIP. The *PARP1* missense variant (rs1136410-G, V762A) is also a significant expression reducing eQTL in whole blood (P = 7.2 × 10^−17^)^14^, and has been recently reported to associate with improved prognosis and survival in MDS^30^. Given the well-established role of *PARP1* in DNA repair^31^, and the approval and application of *PARP1* inhibiting drugs for the treatment of BRCA-positive cancers^32^, it seems plausible that a *PARP1* expression reducing eQTL might antagonize CHIP clone expansion. Furthermore, a recent study used CRISPR screens in zebrafish to identify *PARP1* inhibition as a selective killer of *TET2* mutant hematopoietic stem cells^33^. On the other hand, *PARP1* inhibition is known to cause hematologic toxicity and to increase the risk of treatment related hematologic malignancy^34^. Therefore, further research is needed to test whether *PARP1* inhibition may be appropriate for use in antagonizing the expansion of CHIP clones, and whether any effect is CH-subtype specific.

The *LY75* missense variant (rs78446341-A, P1247L) is located in the extracellular domain of lymphocytic antigen 75, which is also known as DEC-205/CD205, and plays a role in antigenic capture, processing, and presentation^35^. *LY75* is predominantly expressed in hematopoietic derived cells (and particularly dendritic cells)^36^, and its ablation impairs T cell proliferation and response to antigen challenge^37^. The protective association with this variant that we identified appears to be *DNMT3A*-CHIP and mLOY specific, and highlights *LY75* as a potential therapeutic target for the antagonization of *DNMT3A*-CHIP and/or CH.

While most of the phenotypic associations we identified in our cross-sectional analyses were expected associations with hematologic and oncologic traits, the associations we identified with obesity/mass traits are of particular interest. This relationship between mass and CHIP may relate to inflammatory or hormonal signaling, and directions of effect we estimate are consistent with recent findings that *DNMT3A*-CHIP reduces bone mineral density via increases in macrophage-mediated IL20 signaling^38^. The fact the association we report between obesity/mass and CHIP are in opposing directions across CHIP subtypes (e.g. negative in *DNMT3A*-CHIP and positive in *TET2*-CHIP and *ASXL1*-CHIP) suggests that the relationship between CHIP and adiposity is complex, and needs to be investigated further.

Perhaps most unexpectedly, in longitudinal analyses of incident disease, we found that associations between CHIP and CVD and CHIP and myeloid leukemia are more modest than previously reported^1–3^, and that *DNMT3A*-CHIP was not associated with incident lymphoid leukemia. The fact that we found significant associations between blood cancers and *DNMT3A* mutation carriers that also have other CHIP mutations, but not individuals conditioned only on the presence of *DNMT3A* mutations (i.e. agnostic to whether they develop other CHIP mutations prior to developing lymphoid leukemia), helps to isolate the effects on disease risk of *DNMT3A* versus other CHIP mutations. This pattern suggests that hematologic malignancies are predominantly driven by non-*DNMT3A* mutations (and by *TET2*, specifically, among the most recurrently mutated CHIP genes).

A similar pattern was seen for incident CVD, which was consistent with the absence of any relationship between CVD and a *DNMT3A*-CHIP-based polygenic risk score. However, this pattern is not seen across CHIP associations with solid tumors, which we estimate to be driven predominantly by *DNMT3A*. Consistent with this, *DNMT3A*-based polygenic risk scores were associated with multiple cancers and reaffirmed our finding that *DNMT3A*-CHIP is an independent risk factor for lung cancer. Interestingly, in UK Biobank, out of the 3,484 individuals we excluded due to their diagnosis of blood cancer prior to DNA collection and sequencing, 272 (7.8%) had *DNMT3A* mutations, and out of the 40,912 individuals we excluded due to their diagnosis of any cancer prior to DNA collection and sequencing, 2,103 (5.1%) had *DNMT3A* mutations. 18.75% of the *DNMT3A* carriers with blood cancer prior to sequencing had CHIP mutations in multiple genes, whereas only 6.6% of the *DNMT3A* carriers with any cancer prior to sequencing had CHIP mutations in multiple genes. Therefore, the inclusion of such individuals in the analyses done by prior studies, and/or the failure to identify *DNMT3A* carriers as having other CHIP mutations as well, may have led to the misestimation of *DNMT3A* specific risk. On the whole, these results further clarify the role of CHIP mutational subtypes in the development of cancer and CVD, and emphasize the importance of viewing (and potentially treating) different CHIP subtypes as distinct hematologic pre-conditions.

While Bick et al.^13^ found statistical evidence of reduced CVD incidence among CHIP carriers with an *IL6R* coding mutation (rs2228145-C) serving as a genetic proxy for IL-6 inhibition, we did not find any support for this association when extending their analysis from the first 50K exomes in UKB to the full cohort of 450K exomes. The signal identified across the first 50K exomes may result from an ascertainment bias, as those samples were selected to enrich for diagnoses of asthma and individuals with magnetic resonance imaging from the UKB Imaging Study^39^. Alternatively, while the rs2228145-C variant is thought to reflect IL-6 inhibition, and therefore confer protection from heart disease^40^, neither our analysis nor Bick et al. found evidence that rs2228145 carriers are protected from CVD in subjects without CHIP. Therefore, it is possible that this mutation is a poor proxy for IL-6 inhibition, and that direct pharmacological inhibition of IL-6 may still antagonize the interplay between CHIP clone expansion and the onset of CVD.

While this study benefited from having large biobank-scale size and leveraged this to further resolve CH subtypes and broadly assess clinical phenotypes associated with CHIP, limitations include a lack of serial sampling. Such sampling would allow for the evaluation of changes to CHIP clones over time, and future studies that focus on doing such serial analysis at large scale will be able to better estimate CHIP-subtype-specific clonal changes, phenotypic associations, and the risk of CHIP-related clinical outcomes. Such increased data assets would also likely facilitate the identification of additional genes that show recurrent mutation during clonal hematopoiesis, as well as how such mutations relate to one another (i.e. dependency, mutual exclusivity, and temporal order). Nonetheless, we have identified many novel common and rare variant associations with CHIP and other CH phenotypes, which help to set the stage for future functional, mechanistic, and therapeutic studies. On the whole, our analyses emphasize that CHIP is really a composite of somatic mutation driven subtypes, with shared genetic etiology and distinct risk profiles.

## Methods

### Exome sequencing and variant calling

Sample preparation and sequencing were done at the Regeneron Genetics Center as previously described^9,39^. Briefly, sequencing libraries were prepped using genomic DNA samples from the UKB Biobank, followed by multiplexed exome capture and sequencing. Sequencing was performed on the Illumina NovaSeq 6000 platform using S2 (first 50,000 samples) or S4 (all other samples) flow cells. Read mapping, variant calling, and quality control were done according to the SPB protocol described in Van Hout et al.^39^ which included the mapping of reads to the hg38 reference genome with BWA MEM, the identification of small variants with WeCall, and the use of GLnexus to aggregate these files into joint-genotyped, multi-sample VCF files. Depth and allelic valance filters were then applied, and samples were filtered out if they showed disagreement between genetically-determined and reported sex, high rates of heterozygosity/contamination (VBID > 5%), low sequence coverage, or genetically-determined sample duplication.

### Calling CHIP

To call CHIP carrier status, we first used the MuTect2 (GATK v4.1.4.0) somatic caller^41^ to generate a raw callset of somatic mutations across all individuals. This software aims to use mapping quality measures as well as allele frequency information to identify somatic mutations against a background of germline mutations and sequencing errors. We used data generated from gnomAD v2 as the reference source for germline allele frequency^42^. We generated a cohort-specific panel of normals (PON), which MuTect2 uses to estimate per-site beta distribution parameters for use in refining somatic likelihood assignment. Since CHIP is strongly associated with age, we chose 100 random UKB samples aged 40 years old and 622 samples <18 years old samples in GHS to build these cohort-specific PONs. As an initial refinement step, we selected variants occurring within 56 genes that have been recurrently associated with CHIP according to recent reports from the Broad^2^, the TOPMed Consortium^10^, and the Integrative Cancer Genomics (IntOGen) project^43^. We then filtered putative somatic mutations using the functional criteria outlined in Jaiswal et al. 2017^2^. Next, we performed additional QC steps, which consisted of (i) removing multi-allelic somatic calls (ii) applying sequencing depth filters (DP ≥ 20; AD ≥ 3, F1R2 and F2R1 read pair depth ≥ 1), (iii) removing sites flagged as panel of normals by Mutect2 (unless previously reported), (iv) removing indels flagged by the MuTect2 position filter, (v) removing sites within homopolymer runs (a sequence of ≥5 identical bases) if AD < 10 or VAF < 0.08, (vi), removing missense mutations in *CBL* or *TET2* inconsistent with somaticism (i.e., p-value > 0.001 in a binomial test of VAF = 0.5), (vii) removing novel (not previously reported) variants that exhibited characteristics consistent with germline variants or sequencing errors. Given that >90% of mutations belonged to 23 recurrent CHIP-associated genes, we restricted to variants occurring within these genes as a final step to maximize the specificity of our callset. Our final CHIP callset consisted of 29,669 CHIP mutations across 27,331 unique individuals from UK Biobank, and 14,766 CHIP mutations across 12,877 unique individuals from GHS. Variant allele fraction (VAF) was calculated using AD/DP.

### Defining CHIP and mosaic phenotypes

CHIP phenotypes were derived based on our mutation callset, whereas mosaic chromosomal alteration (mCA) phenotypes were derived based on previously published mCA calls from the UKB Biobank^4,6,7^. First, we used ICD codes to exclude 3,596 samples from UK Biobank and 1,222 samples from GHS that had a diagnosis of blood cancer prior to sample collection. We also excluded 13,004 individuals from GHS whose DNA samples were collected from Saliva as opposed to blood. We then defined multiple CHIP and mosaic phenotypes based on the whether carriers did (inclusive) or did not (exclusive) have other somatic phenotypes. For example, individuals with at least one CHIP mutation in our callset were defined as carriers for a CHIP_inclusive phenotype, whereas anyone with a CHIP mutation as well as an identified mCA was removed from this inclusive phenotype in order to define a CHIP_exclusive phenotype (21,587 cases and 364,072 controls). Our association analysis with CHIP used this CHIP_inclusive phenotype, which included 26,734 cases and 364,073 controls in UK Biobank, and 12,480 cases and 148,849 controls in GHS. We defined mLOY carriers as male individuals with a Y chromosome mCA in the UK Biobank mCA callset that had copy change status of loss or unknown, mLOX as individuals with an X chromosome mCA in the UK Biobank mCA callset that had copy change status of loss or unknown, and mCAaut carriers as individuals with autosomal mCAs. We then refined these inclusive phenotypes to define exclusive versions, with mLOY_exclusive consisting of carriers with no X chromosome or autosomal mCAs (36,187 cases and 151,161 controls), mLOX_exclusive consisting of carriers with no Y chromosome or autosomal mCAs (10,743 cases and 364,072 controls), and mCAaut_exclusive consisting of carriers with no Y or X chromosomal alterations of any kind (11,154 cases and 364,072 controls). These exclusive phenotypes were used for all analyses comparing CHIP with mosaic phenotypes, as this approach facilitated the generation of four non-overlapping phenotypes (i.e. CHIP, mLOY, mLOX, and mCAaut) that could be compared and contrasted. We also defined CHIP gene-specific phenotypes by choosing carriers as those with mutations in our callset from a specific gene and no mutations in any other of the 23 CHIP genes defining our callset. For example, CHIP-*DNMT3A* carriers were those with ≥ 1 somatic mutations in our callset within the *DNMT3A* gene, and no mutations in our callset in any of the other 23 CHIP genes we used for our final callset definition.

### Genetic association analyses

To perform genetic association analyses, we used the genome-wide regression approach implemented in REGENIE^44^, as described in Backman et al.^9^. Briefly, regressions were run separately for data derived from exome-sequencing as well as data derived from genetic imputation using TOPMed^45^, and results were combined across these data sources for downstream analysis. Step 1 of REGENIE uses genetic data to predict individual values for the trait of interest (i.e. a PRS), which is then used as a covariate in step 2 to adjust for population structure and other potential confounding. For step 1, we used variants from array data with a minor allele frequency (MAF) >1%, <10% missingness, Hardy-Weinberg equilibrium test *P*-value>10^−15^ and linkage disequilibrium (LD) pruning (1000 variant windows, 100 variant sliding windows and *r*^2^<0.9), and excluded any variants with high inter-chromosomal LD, in the major histocompatibility (MHC) region, or in regions of low complexity. For association analyses in step 2 of REGENIE, we used age, age^2^, sex, and age-by-sex, 10 ancestry-informative principal components (PCs) as covariates. For analyses involving exome data, we also included as covariates an indicator variable representing exome sequencing batch, and 20 PCs derived from the analysis of rare exomic variants (MAF between 2.6×10^−5^ and 0.01). Significance cutoffs were set according to the power calculations and logic outlined by Backmen et al.^9^. Results were visualized and processed using an in-house version of the FUMA software^46^.

Associations for CHIP subtypes *JAK2* and *SRSF2* were not replicated in the GHS cohort as we had too few samples for genome-wide model convergence. Association analyses were performed separately for different continental ancestries defined based on the array data, as described in Backman et al.^9^.

### Pairwise mutational analyses

Pairwise mutational enrichments across our UK Biobank and GHS CHIP callsets were calculated using Fishers’s Exact test among carriers with >=2 somatic CHIP mutations. Results were qualitatively similar when restricting the age range. For individuals with more than two CHIP mutations, we made the simplifying assumption of considering each mutation pair independently in our tested counts.

### Genetic comparisons between CHIP subtypes

For pairwise comparisons between CHIP gene mutation subtypes, we used the union set of index SNPs (i.e. independent signals in genome-wide significant loci) from all of our CHIP and CHIP gene subtype associations. This resulted in 91 variants, which we used to compare effect sizes estimates between CHIP subtype pairs. Associations were estimated using linear regression, with the estimated effect size of variants in trait one as the dependent variable and the estimated effect size of variants in trait two as the independent variable. We also used the ggplot2 package in R to visualize all CHIP subtype comparisons together. Genetic correlations and trait heritability estimates were calculated using ldsc version 1.0.1 with annotation input version 2.2^21^.

### Pairwise comparisons between CHIP and mosaic phenotypes

For pairwise comparisons between CHIP and mLOY, mLOX, and mCAaut, we used the union set of index SNPs from all of our CHIP, CHIP gene subtype, and mosaic associations. This resulted in 341 variants, which we used to compare effect sizes estimates between phenotypic pairs. Pairwise associations were estimated using linear regression, with the estimated effect size of variants in trait one as the dependent variable and the estimated effect size of variants in trait two as the independent variable. Genetic correlations and trait heritability estimates were calculated using ldsc version 1.0.1 with annotation input version 2.2^21^.

### Phenotypic associations with CHIP and CHIP-subtypes

To test for known as well as potentially novel associations, we used Regenie^47^ to perform Firth-corrected tests for association between our 23 CHIP gene-specific phenotypes and 4,871 traits (2,459 binary traits and 2,421 quantitative traits) from the UK Biobank (V5). To do this, we coded each CHIP gene-specific phenotype as 1 if an individual had any somatic CHIP mutation in the gene and 0 otherwise, and formatted these binary codings as though they were genotypes in order to run regressions with Regenie. Regressions were run as described previously, with age, sex, and genetic PCs as covariates^9^. After filtering out association tests where the total number of cases that were also somatic carries was < 5 and/or those that did not converge, we were left with 81,814 total association tests (Table S40). Quantitative traits were transformed using a reverse inverse normalized transformation (RINT), and so effect size estimates from these associations are in units of standard deviation. To visualize high-level phenotypic patterns across these CHIP gene-specific phenotypes (Figure 5), we categorized phenotypes by disease group^9^, and calculated the proportion of phenotypes per disease group per gene that were associated at a P <= 0.05 alpha level (uncorrected). To visualize the most significant of these associations we plotted effect sizes (Figure S7) by disease category for all associations with P <= 1e-5 (0.05/5000).

### Longitudinal analysis of hematological and oncological risk within individuals with CHIP

We performed longitudinal survival analyses using cox proportional hazard models (coxph function) as implemented in the survival r package. Given that CHIP is strongly correlated with age, and that time to event intervals will reflect older age periods for CHIP carriers, we set up models with age as the time variable, with interval censoring using age at first assessment and age at event or censoring. Individuals with follow-up time in excess of 13.5 years (3% of the dataset) were censored due to departures from the proportional hazards model. This allows for an implicit adjustment for age within the proportional hazard models. Analyses were performed on individuals of European ancestral background. All models included 10 genetically determined European-specific PCs as covariates, and all analyses excluded individuals genetically determined to be 3^rd^ degree relatives or closer. We used a variety of CHIP codings as variables in our models to test for potential differences between high/low VAF CHIP and/or CHIP subtypes. These included (i) any individuals with a CHIP variant in our callset (ii) CHIP carriers with >= 1 CHIP mutation with VAF >= 0.10 (iii) carriers of CHIP mutations in *DNMT3A* (iv) carriers of CHIP mutations in *DNMT3A* with VAF >= 0.1 (v) carriers of CHIP mutations in *TET2* (vi) carriers of CHIP mutations in *TET2* with VAF >= 0.1 (vii) carriers of CHIP mutations in *ASXL1* (viii) carriers of CHIP mutations in *ASXL1* with VAF >= 0.1 (ix) carriers of any *DNMT3A* or *TET2* mutations (x) strict CHIP mutation definitions that subset our callset to mutations specified in previous publications (e.g. Jaiswal et al.^2^) (xi) carriers of *DNMT3A* mutations with at least one additional CHIP mutation and another CHIP gene. Note that the CHIP gene specific codings just described vary from the phenotypic coding definitions used in our GWAS/ExWAS, which required carriers to have mutations in the specified CHIP gene and no mutations in any other CHIP genes. Since mutational exclusivity in addition to the VAF >= 0.1 threshold would have required an individual to have a high VAF CHIP gene mutation but also not have any other mutations, which is not realistic and also significantly lowers sample size, we chose this adjusted definition for these longitudinal analyses of disease incidence. For the composite phenotypes described below, we used a combination of self-reported data, general practitioner and hospital records and procedures codes (OPCS4) to define prevalent disease (i.e. disease occurring before sample collection, which was used to exclude samples from longitudinal analysis of incident disease). Incident disease was then determined using the codings described below.

For the CVD analyses, we included sex, LDL, HDL, pack years, smoking status (current vs former), BMI, essential primary hypertension, and type 2 diabetes mellitus as covariates. The CVD composite outcome was derived from the health codes listed in Table S41. The results we reported used a composite of myocardial infarction (MI), coronary artery bypass graft (CABG), percutaneous coronary intervention (PCI), and coronary artery disease (CAD), and also included death from any of these events. Results were the same when our composite included ischemic stroke (ISCH.TR), as well as when we repeated analyses with a stricter subset of recurrent CHIP mutations derived from Jaiswal et al.^2^ or with variants only found in *DNMT3A* or *TET2*. We also excluded samples with any diagnosis of malignant blood cancer prior to sequencing (n = 3,596). Missing LDL and HDL values were median imputed, and individuals on cholesterol medication had their raw LDL values increased by a factor of 1/0.68, similar to Bick et al.^13^ *IL6R* missense variant (rs2228145-C) genotypes were modeled dominantly (coded as 1 for carriers of any allele and 0 otherwise), and we modeled the effect of this allele in additive, interactive, and CHIP status stratified proportional hazard models. Models considering only the initial 50k UK Biobank individuals restricted to the samples reported by Bick et al.^13^. For visualization, we estimated base (Kaplan Meyer) survival curves (i.e. no covariates) using the survfit function in the aforementioned Survival package, and made plots using the ggsurvplot function from the survminer package.

For models of cancers and overall survival risk tested using all CHIP carriers, high VAF (VAF >= 0.1) CHIP carriers, and carriers of specific CHIP gene mutations, we used unrelated European samples that did not have any cancer diagnoses prior to sample collection (N = 360, 051 after the removal of 33,816 samples with a prior diagnosis of cancer). Results were qualitatively the same when repeating these analyses without excluding samples that had a diagnosis of any malignant cancer prior to sample collection date. Cancer phenotype definitions were derived from medical records indicating the following ICD10 codes: C81-C96, D46, D47.1, D47.3, D47.4 for blood cancers, C81-C86, C91 for lymphoid cancers, C90, C92, C94.4, C94.6, D45, D46, D47.1, D47.3, D47.4 for myeloid cancers, C50 for breast cancers, C34 for lung cancers, C61 for prostate cancers, and C18 for colon cancers. For blood cancers, we also included cases that self-reported having leukemia, lymphoma, or multiple myeloma. These models were implemented with the same covariates and in the same fashion as described above, although models estimating risk for sex specific cancers (i.e. prostate and breast) restricted to individuals of the relevant sex and did not adjust for sex as a covariate. For smoking stratified modeling of blood and lung cancer, we used a stricter definition of smoking (ever vs never) and included pack years as a covariate in models testing risk among smokers.

### Polygenic risk scores

Polygenic risk scores were calculated with Plink^47^ using a clumping and thresholding approach. First, summary statistics from a genome-wide association of DNMT3A-CHIP in UKB were filtered to only include variants with a minor allele frequency >= 0.01. We then performed clumping with plink1.9 using the 1KG EUR data as an LD reference, 5e-8 for both p-value thresholds, and an R^2^ threshold of 0.2. This resulted in 72 variants, which we used along with plink2.0 to generate polygenic risk scores in the UKB cohort. We then performed association tests using logistic regression, with binary phenotypes of interest as the dependent variable, this polygenic risk score as the independent variable of interest, and age, sex, smoking status (ever vs never), and 10 genetic PCs as covariates. Outcomes of interest were those from our longitudinal analyses, but updated to include prevalent disease cases (as a PRS serves as a genetic instrument whose exposure can be considered from birth). We also modeled carrier status for each of the top-8 CHIP subtypes as outcomes.

## Figure Legends

**Figure S1.**
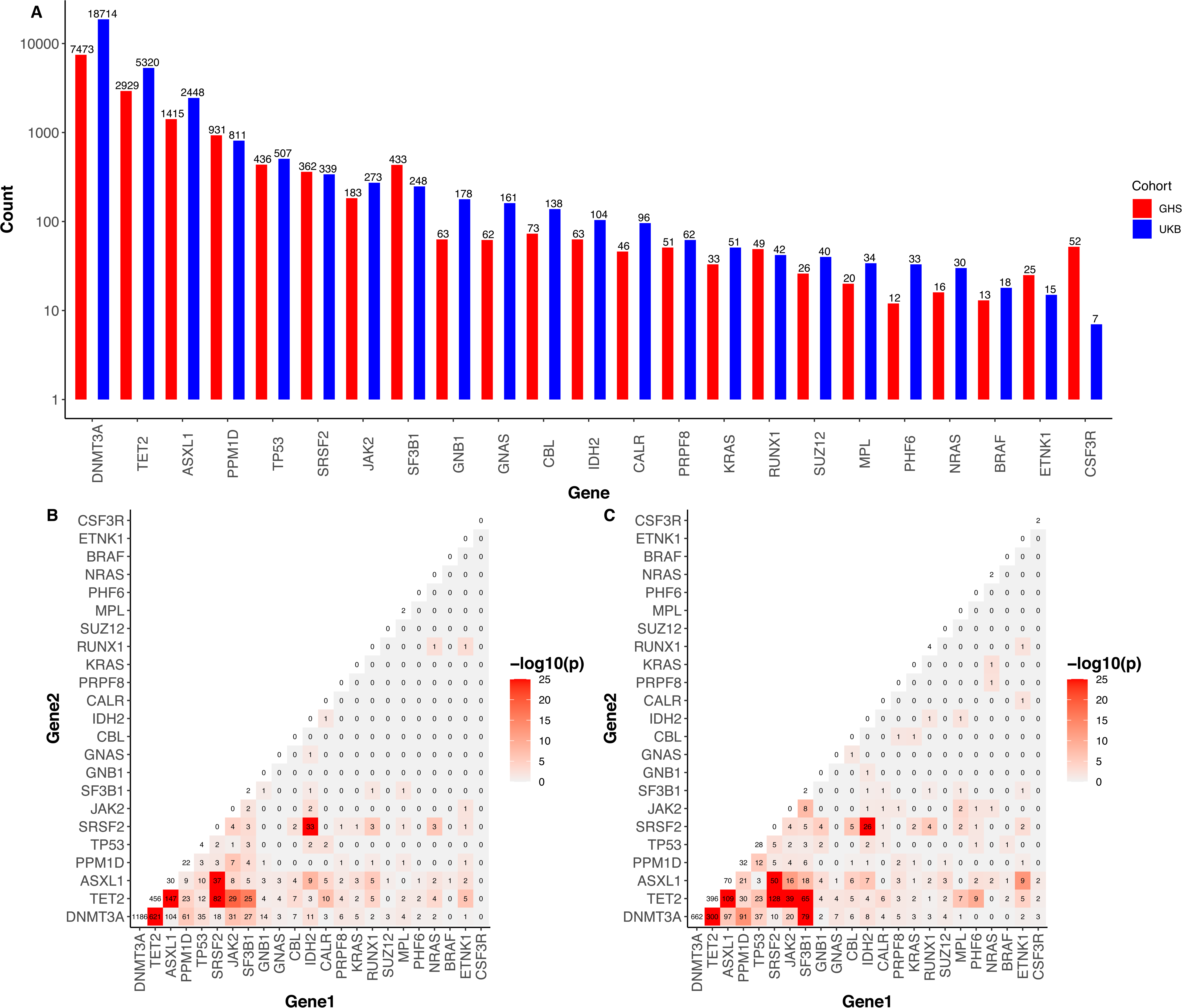
Count Distribution and Pairwise Enrichments of Clonal Hematopoiesis of Indeterminate Potential (CHIP) Gene Mutations. A. Total number of individuals with mutations (y axis, log_10_ scale) in each of the 23 genes that were used to determine CHIP status across the UKB (blue) and GHS (red) CHIP callsets. B-C. Pairwise mutation counts across the UKB (B) and DiscoverEHR (C) callsets across individuals with at least two identified CHIP mutations. The color scale reflects the significance of the P-value for enrichment of the presence of pairs of gene mutations as determined by Fisher’s exact test. P-values are –log_10_ transformed, and values > 25 were set to the color corresponding to 25 for visualization (see Table S1 for complete enrichment results).

**Figure S2.**
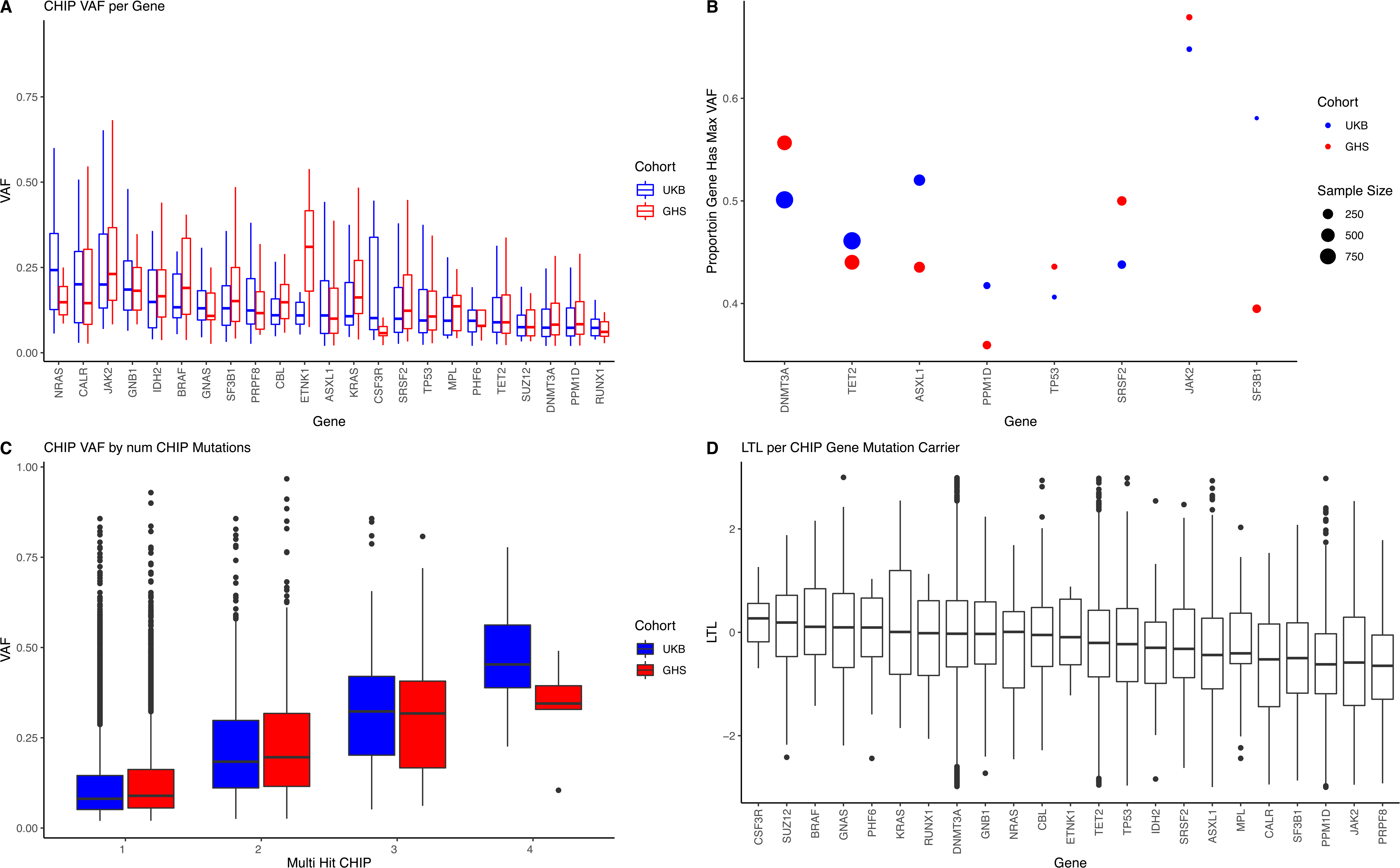
Variant Allele Fraction (VAF) and Leukocyte Telomere Length (LTL) by Specific CHIP Gene Carrier Status. A. Distributions of variant allele fraction (VAF, y axis) are shown for mutations within each of the 23 CHIP genes in our callset across individuals with only a single CHIP mutation from UKB (blue) and DiscoverEHR (red). Canonical CHIP genes generally had low VAFs compared with mutations more canonically associated with leukemias. B. Among individuals with multiple CHIP mutations, the proportion of times each CHIP gene mutation was the one with the highest VAF is shown (y axis) for the top 8 CHIP genes. Higher proportions suggest that those CHIP genes represent either the driver mutation for CHIP or presence in the largest CHIP clone. C. The maximum VAF among all CHIP mutations is higher in individuals with increasing numbers of CHIP mutations. D. Distributions of telomere length (LTL) are shown across 23 CHIP gene mutation carriers in our callset.

**Figure S3.**
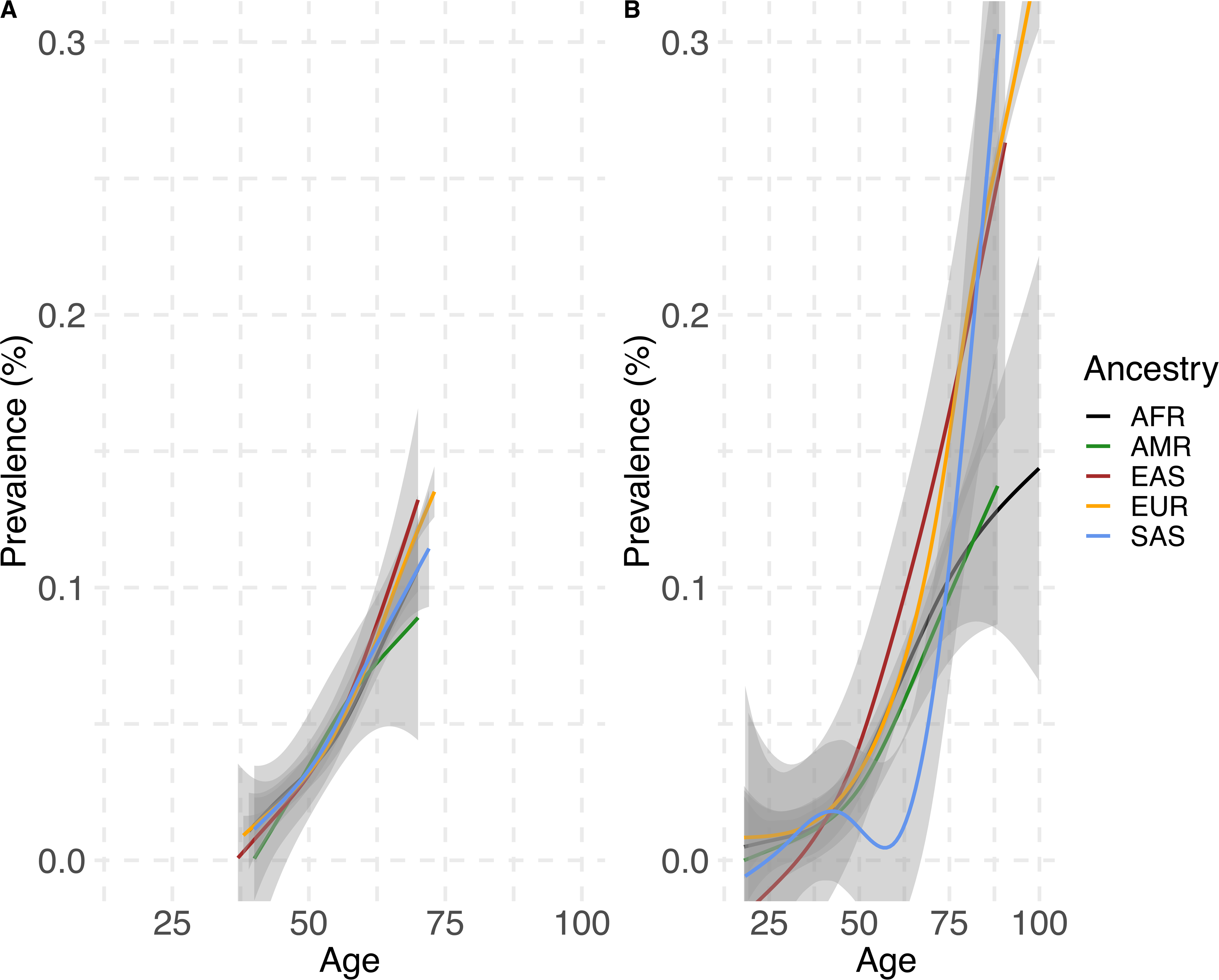
CHIP Prevalence Across Ancestries. Splines showing the prevalence of CHIP mutations by age across different continental ancestries in the UKB and GHS cohorts.

**Figure S4.**
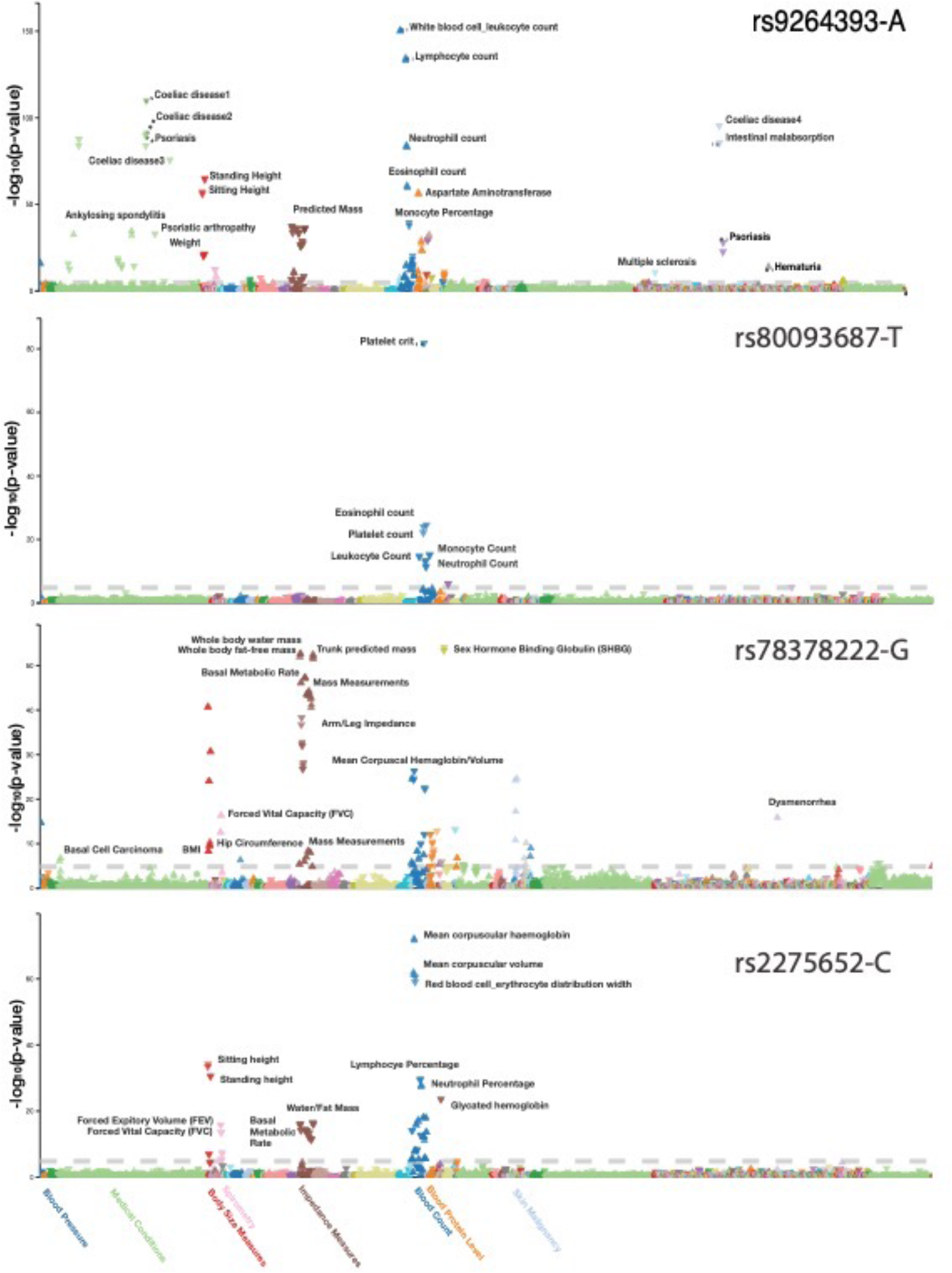
Results From a Phenome-Wide Association Analysis. Results from a phenome-wide association analysis are shown for four CHIP-associated index SNPs from our GWAS. These depict associations with blood, body mass, and auto-immune traits, and are representative of the traits found to be most significantly associated with our CHIP-associated loci.

**Figure S5.**
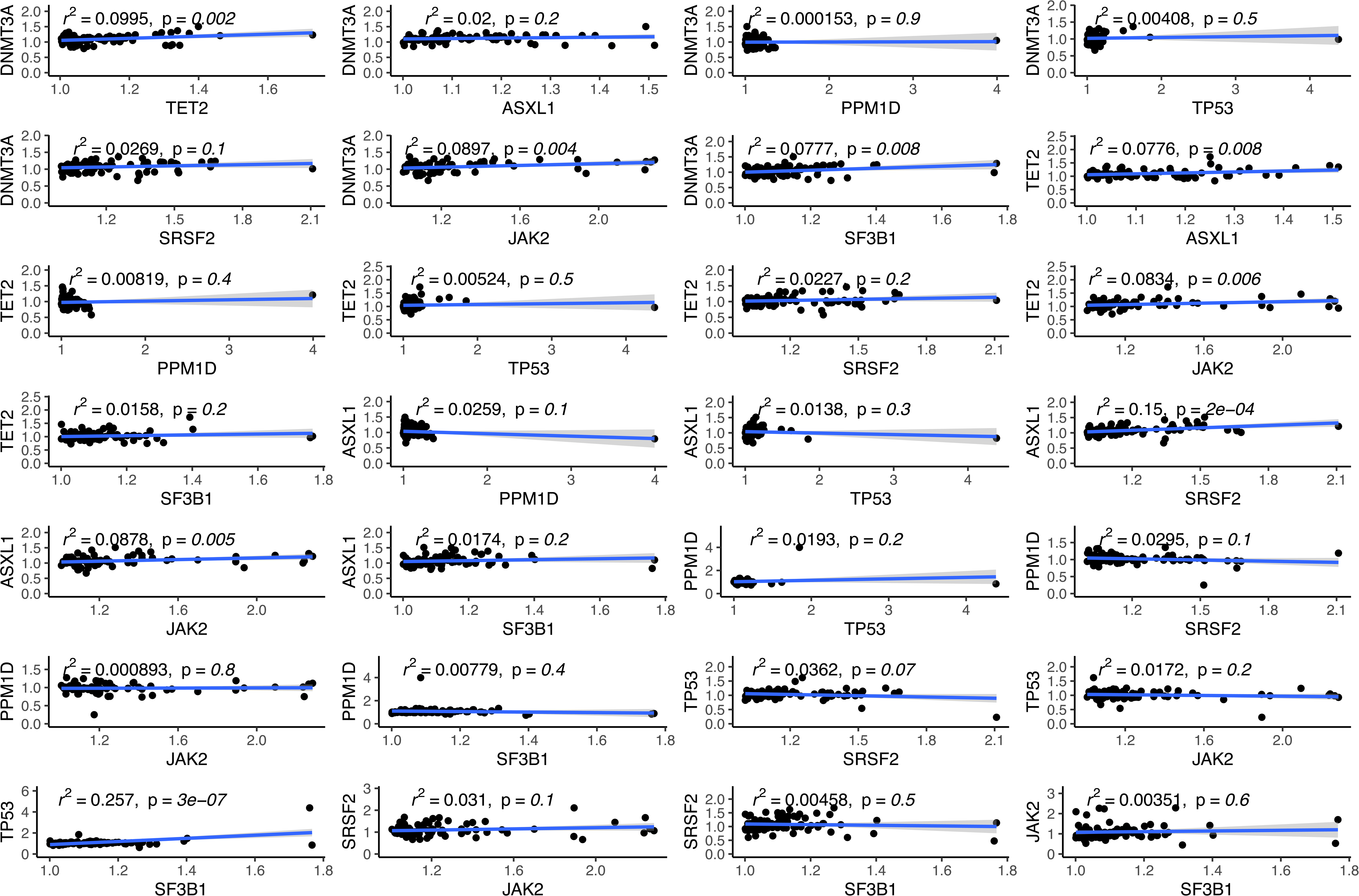
Pairwise Index SNP Effect Size Comparisons Across CHIP Gene Subtypes. Using results from CHIP-gene-specific association analyses, effect sizes of index SNP are compared in pairwise fashion, and linear relationships are evaluated using linear regression.

**Figure S6.**
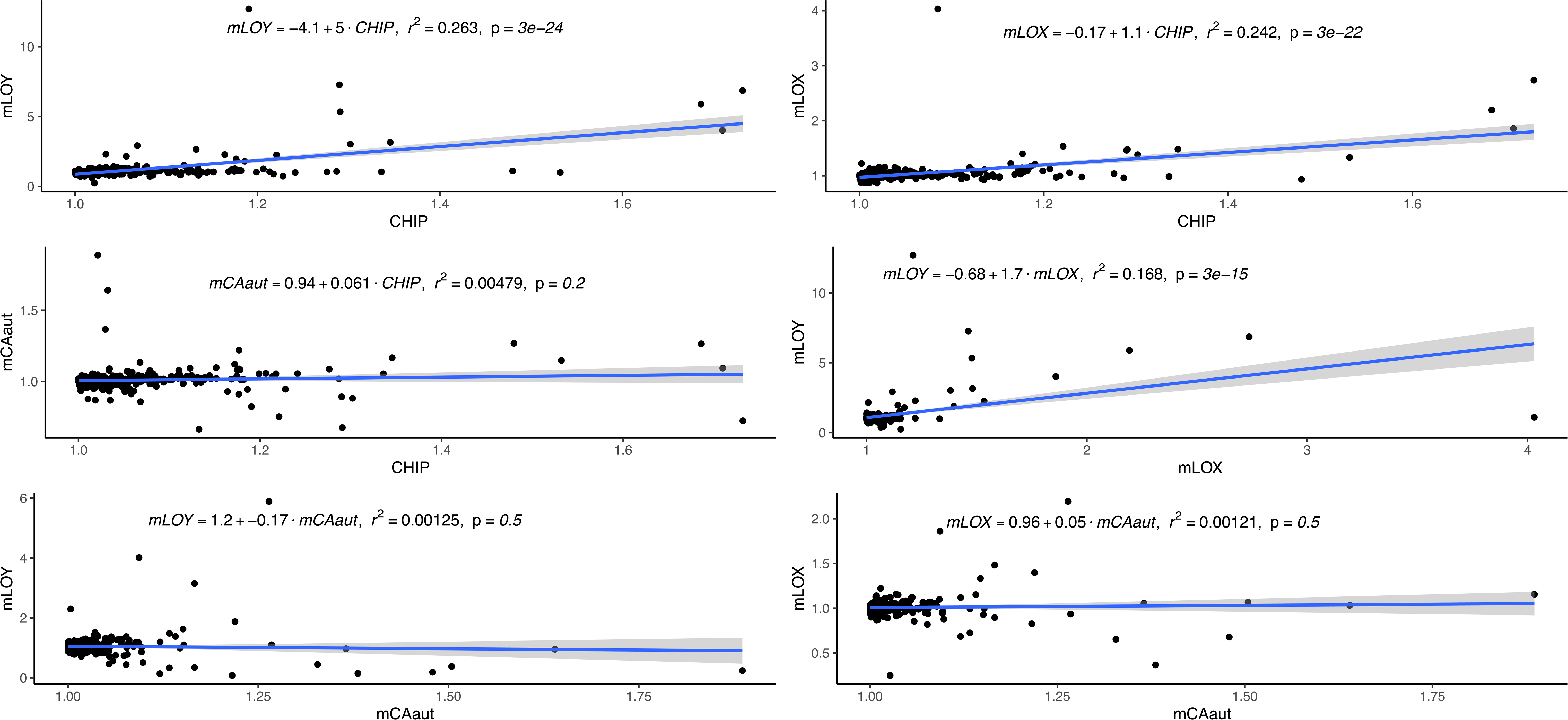
Pairwise Index SNP Effect Size Comparisons Between CHIP and mCA Phenotypes. Using results from exclusive CHIP and mCA association analyses, effect sizes of index SNP are compared in pairwise fashion, and linear relationships are evaluated using linear regression.

**Figure S7.**
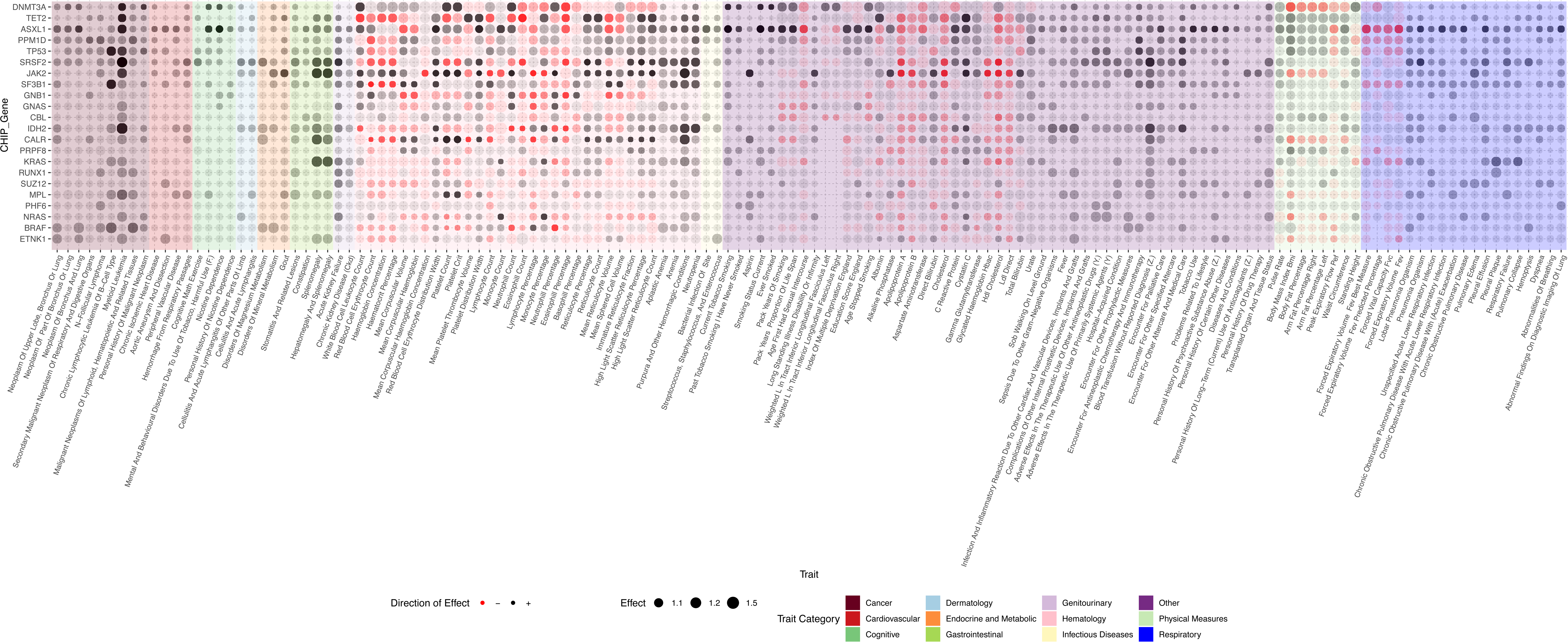
Comparison of Associations Across Phenotypes Associated with CHIP. Association results are visualized across all traits (x-axis) that are associated (P <= 1e-5) with at least one CHIP gene subtype (y-axis). Dot color represents effect size direction, and dot size is scaled to the log10 value of the effect size. Background color represents trait category, and is the same as in Figure 5, although the alpha value is reduced so that the effect size dots can be readily seen.

**Figure S8.**
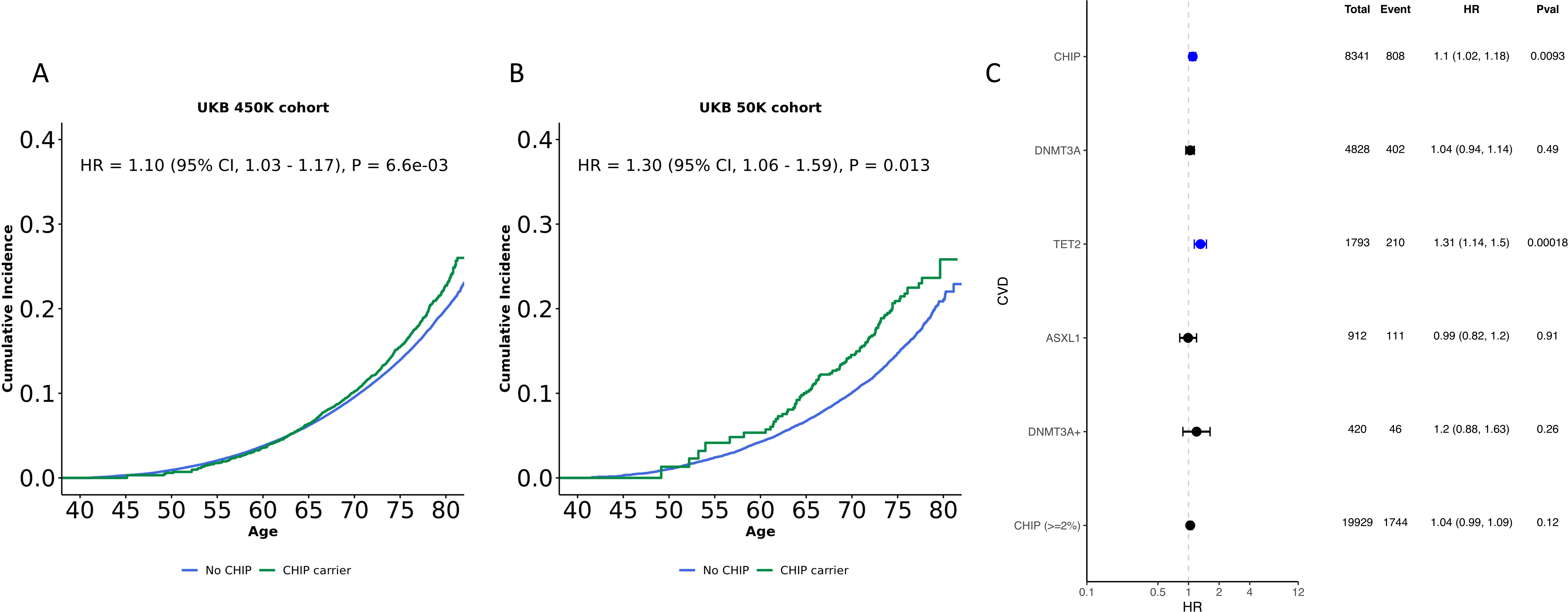
CVD Incidence among CHIP carriers in UKB 50K vs UKB 450K. Survival curves are drawn showing the increased risk of CVD for CHIP carriers, compared to individuals without CHIP. In the first 50,000 individuals from UKB (A), this risk is higher (OR = 1.30) than in the full cohort (B, OR = 1.10). Breaking down CVD risk according to CHIP gene mutation (C) suggests that risk is driven by TET2 mutations (OR = 1.31, P = 1.80 * 10^−4^).

## Study Approval

*UK Biobank study:* ethical approval for the UK Biobank was previously obtained from the North West Centre for Research Ethics Committee (11/NW/0382). The work described herein was approved by UK Biobank under application number 26041. *GHS study:* approval for DiscovEHR analyses was provided by the Geisinger Health System Institutional Review Board under project number 2006-0258.

## Author Contributions

MDK, AD, SO, SS, NB, DL, and HMK performed bioinformatic and statistical genetic analysis. SO, KW, CG, JM, WS, JGR, and HMK contributed to data engineering and bioinformatic pipeline development. JDO developed and oversaw exome sequencing efforts. MDK, AD, SO, JH, MVM, YH, KW, CG, AL, JAK, RW, MGL, MJ, DJG, LAL, MNC, GSA, MARF, RQ, GT, CP, ARS, WS, JGR, JDO, JM, HMK, AB, GRA, EJ contributed to experimental design, interpretation of results, and genetic program development. MDK and EJ drafted the manuscript, with significant input from AD, SO, MVM, GT, HMK, AB, and GRA. All authors reviewed and approved the final version of the manuscript.

## Data Availability

Dervied data (e.g. CHIP calls) produced in the present study are available upon reasonable request to the authors

